# Multivariate variance components analysis uncovers genetic architecture of brain isoform expression and novel psychiatric disease mechanisms

**DOI:** 10.1101/2022.10.18.22281204

**Authors:** Minsoo Kim, Daniel D. Vo, Connor T. Jops, Cindy Wen, Ashok Patowary, Arjun Bhattacharya, Chloe X. Yap, Hua Zhou, Michael J. Gandal

## Abstract

Multivariate variance components linear mixed models are fundamental statistical models in quantitative genetics, widely used to quantify SNP-based heritability (*h*^2^_SNP_) and genetic correlation (*r*_g_) across complex traits. However, maximum likelihood estimation of multivariate variance components models remains numerically challenging when the number of traits and variance components are both greater than two. To address this critical gap, here we introduce a novel statistical method for fitting multivariate variance components models. This method improves on existing methods by allowing for arbitrary number of traits and/or variance components. We illustrate the utility of our method by characterizing for the first time the genetic architecture of isoform expression in the human brain, modeling up to 23 isoforms jointly across ∼900 individuals within PsychENCODE. We find a significant proportion of isoforms to be under genetic control (17,721 of 93,293 isoforms) with substantial shared genetic effects among local (or *cis*-) relative to distal (or *trans*-) genetic variants (median *r*_g,cis_ and *r*_g,trans_ = 0.31 and 0.06). Importantly, we find that 11.6% of brain-expressed genes (2,900 genes) are heritable only at the isoform-level. Integrating these isoform-specific genetic signals with psychiatric GWAS signals uncovers previously hidden psychiatric disease mechanisms. Specifically, we highlight reduced expression of a specific *XRN2* isoform as the underlying driver of the strongest GWAS signal for autism spectrum disorder. Overall, our method for fitting multivariate variance components models is flexible, widely applicable, and is implemented in the Julia programming language and available online.

## Introduction

The genetic contribution to phenotypes of interest (i.e. SNP-based heritability or *h*^2^_SNP_) and the extent to which they are shared (i.e. genetic correlation or *r*_g_) can be quantified through variance components linear mixed models. The simplest univariate variance components model assumes two variance components, one of which captures the aggregate genome-wide genetic effects^1^. However, this model is likely misspecified for either gene or isoform expression, since SNPs in the vicinity of a gene (herein referred to as *cis*-SNPs) tend to exert stronger effects on its expression than distal SNPs (i.e. *trans*-SNPs)^2^. Similarly, the simplest multivariate variance components model looks at phenotypes pairwise and assumes two variance components, one of which captures the aggregate degree of pleiotropy among genome-wide SNPs^3,4^. Again, this model is likely inadequate for isoform-level expression, since the degree of genetic correlation can differ among different sets of SNPs (e.g. *cis*- and *trans*-SNPs).

Unfortunately, maximum likelihood (ML) and restricted maximum likelihood (REML) estimation of the most general form of multivariate variance components models with more than two phenotypes and more than two variance components still poses a significant numerical and computational challenge. Existing methods lack flexibility by modelling at most two phenotypes with multiple variance components^5,6^ or multiple phenotypes with two variance components^7,8^. To address this critical gap, we implemented the minorization-maximization (MM) algorithm^9^ for ML and REML estimation in multivariate variance components linear mixed models^10^, using the Julia programming language^11^. We refer to the Methods for a comprehensive summary of optimization methods for tackling variance components models, but we note here that the major advantages of the MM algorithm include numerical stability, fast convergence, and graceful adaptation to the positive semidefiniteness constraint of variance components parameters^10^.

To demonstrate the utility of our method, we modelled constituent isoforms of a given gene jointly in the PsychENCODE dataset^12,13^, partitioning genetic variance and covariance among *cis*- and *trans*-SNPs. We find that genetic variants local to the gene or isoform in question have a sparse but large effect on expression, whereas distal genetic variants have individually small but collectively large effects on expression. We further show that there are substantial shared genetic influences among *cis*-SNPs compared to *trans*-SNPs for isoform expression. Importantly, we find isoform-level analyses to lead to discovery of many more genetic signals than gene-level analyses. Some of these isoform-specific genetic signals were significantly associated with increased risk for psychiatric disorders such as autism spectrum disorder^14^. These findings suggest that isoform-resolution analyses have the potential to uncover novel disease genes and aid in interpreting GWAS results. Overall, we present a comprehensive dissection of genetic influences on brain gene and isoform expression and their relation to brain-related disorders.

## Results

### MM algorithm for multivariate variance components linear mixed models

The multivariate variance components linear mixed model with *n* ×*d* response matrix **Y**, *n* ×*d* predictor matrix **X**, and *m* known *n* × *n* positive semidefinite matrices (**V**_1_,…, **V**_*m*_) (e.g. genetic relationship matrices) assumes vec **Y** ∼ *N* (vec(**XB**), **Ω**), where

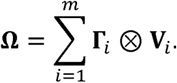

The parameters of the model include *p* × *d* mean effects **B** and *d* × *d* variance components (**Γ**_1_, …, **Γ**_*m*_) that are positive semidefinite. vec **Y** creates an *nd* × 1 vector from **Y** by stacking its columns and ⊗ denotes the Kronecker product. The univariate model is subsumed under the multivariate model when *d* = 1. Under this model, when there are no mean effects or there is only the intercept term, phenotypic variance and covariance can be decomposed into the sum of variance components parameters (Methods).

As there are no closed-form solutions to ML estimation, variance components are usually estimated using an iterative algorithm such as Fisher scoring and expectation-maximization (EM) algorithms. Here, we pursue ML estimation with the MM algorithm^10^ that alternatively updates **B** and **Γ**_*i*_ ‘s. In each iteration, the MM updates are

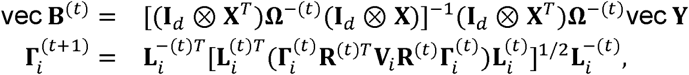

where 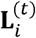 is the Cholesky factor of 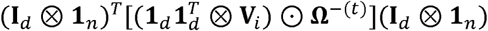 and **R**^(*t*)^ is the *n* × *d* matrix such that vec **R**^(*t*) =^ **Ω**^−(*t*)^vec (**Y** −**XB** ^(*t*)^). ⊙ denotes the Hadamard product. For REML estimation, the updates are basically of the same form. Compared to other iterative algorithms, the MM algorithm is numerically stable, converges fast, and gracefully respects the nonnegativity (or positive semidefinite) constraint of variance components. In the simplest model where there are two variance components, repeated matrix inversion can be avoided by the generalized eigenvalue decomposition of the two kernel matrices^10,15,16^. When there are more than two variance components, a matrix inversion is inevitable in each update, so the MM algorithm is not scalable to biobank-level data, and we recommend applying this method for datasets of size up to *n* × *d =* 50000.

Standard errors for our parameter estimates were subsequently calculated using the Fisher information matrix (Methods), where

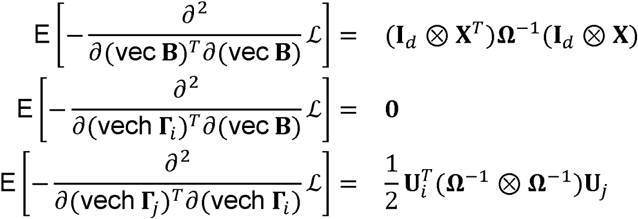

and 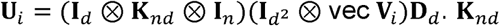 is the *nd* × *nd* commutation matrix and **D**_*d*_ the *d*^2^ × *d*(*d* +1)/2 duplication matrix. vech **Γ**_***i***_ creates an *d*(*d* +1)/2 × 1 vector from **Γ**_*i*_ by stacking its lower triangular part.

### Overview of *h*^2^_SNP_ and *r*_g_ analyses for brain gene and isoform expression

In the present study, for all variance components models, whether be it univariate, pairwise bivariate, or multivariate, we specified three variance components that include *cis*- and *trans*-SNP effects, and residual effects. This model specification corresponds to an assumption that *cis*- and *trans*-SNP effects are realized from different distributions of effect sizes. Hereafter, we refer to the variance components parameters corresponding to either *cis*- or *trans*-SNP effects as genetic variances and genetic covariances. We defined *cis*-SNPs as those within ±1 Mb window of gene start and gene end sites and *trans*-SNPs as all other SNPs (**Figure 1**). Indeed, other definitions are possible, and we tested varying windows as part of sensitivity analyses.

**Figure 1:**
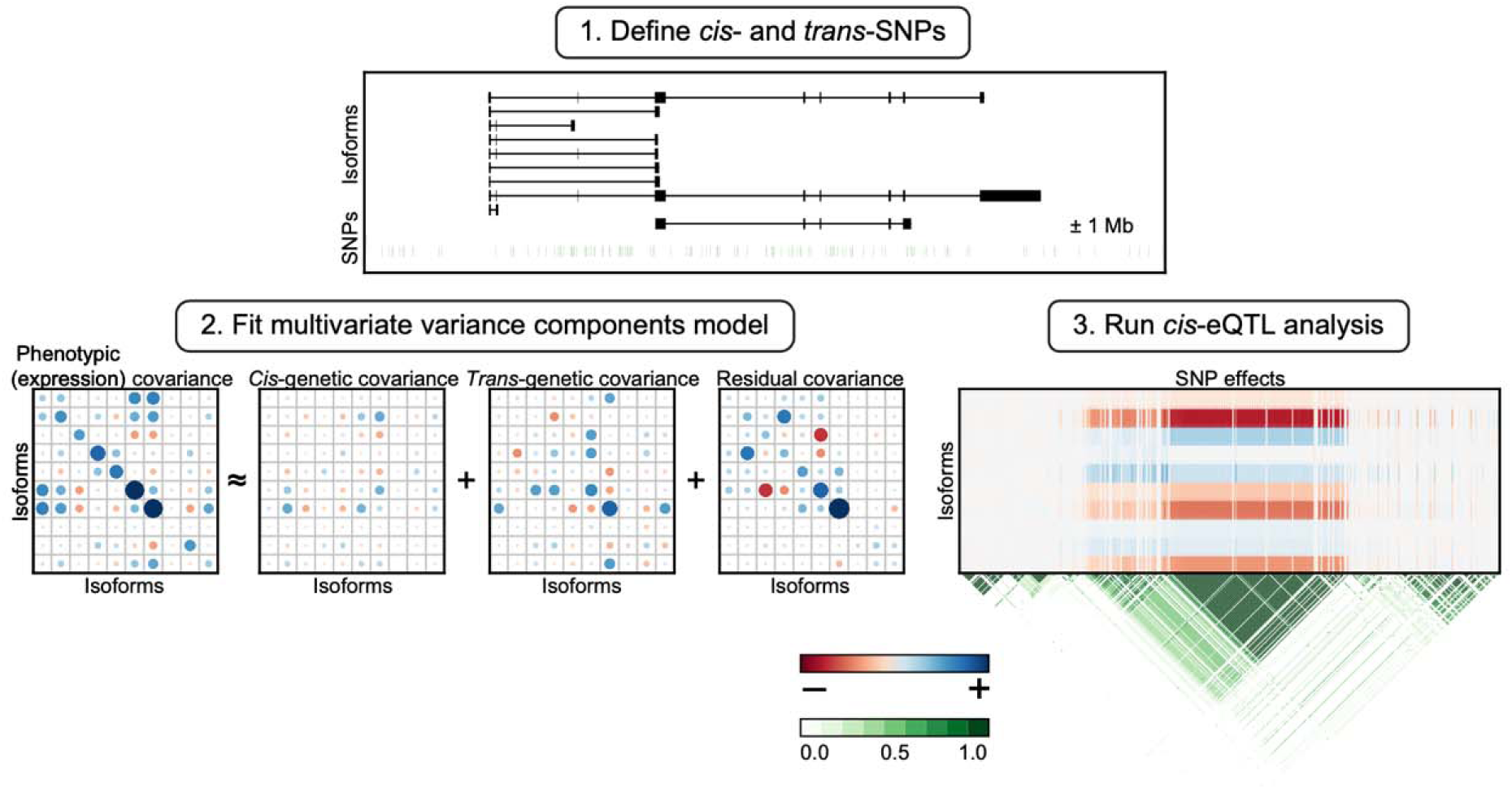
Overview of isoform-centric *h*^2^_SNP_, *r*_*g*_, and *cis*-eQTL analyses. For a given gene and its constituent isoforms, *cis*-SNPs are defined as SNPs within ±1 Mb window of (collapsed) gene start and end sites, while *trans*-SNPs are defined as all other SNPs. Expression variances and covariances are partitioned among *cis*- and *trans*-SNPs for each gene and its constituent isoforms by fitting univariate, pairwise bivariate, and multivariate variance components linear mixed models^10^. The same sets of *cis*-SNPs are used for *cis-*eQTL analyses^18^, results of which are compared to heritability and genetic correlation analyses. This schematic figure is based on the real data for ten isoforms belonging to the *KLHL24* gene.

We present estimates from the MM algorithm and REML estimation, unless otherwise stated. The standard errors for variance components estimates were calculated from the Fisher information matrix. Inference on variance components parameters in the univariate case was done using a variation of the likelihood ratio test (LRT)^5,17^ (Methods). In the multivariate setting, inference on the off-diagonal elements of the variance components parameters was done using the Wald test (Methods).

We first estimated *h*^2^_SNP_ and assessed its significance by fitting the univariate variance components model. Since we specified two separate variance components parameters for *cis*- and *trans*-SNPs, *h*^2^_SNP_ could be decomposed into *h*^2^_cis_ and *h*^2^_trans_ (i.e. *h*^2^_SNP_ = *h*^2^_cis_+ *h*^2^_trans_). To avoid estimating too many parameters in multivariate models, for each gene, we focused on isoforms with significant *h*^2^_SNP_ at *P* < 0.05 in the univariate model. Since we partitioned expression covariances among *cis*- and *trans*-SNPs, and residual effects, there were two genetic correlation parameters (*r*_g_’s) that correspond to aggregate degree of pleiotropy among *cis*-SNPs (*r*_g,cis_) and aggregate degree of pleiotropy among *trans*-SNPs (*r*_g,trans_) (**Figure 1**). There was also residual correlation (*r*_e_) which captures correlation due to biological and technical factors such as shared gene regulation and measurement error (Methods). It is important to note that *r*_g_ is not the sum of *r*_g,cis_ and *r*_g,trans_, since these parameters are on a normalized scale. Finally, we conducted *cis*-eQTL analyses^18^ in parallel with the same set of *cis*-SNPs to validate *h*^2^_SNP_ results as a positive control and to link isoform-specific signals with GWAS signals (**Figure 1**).

### Polygenicity of brain gene and isoform expression

We started with previously harmonized genotype array and frontal cortex RNA-seq data from PsychENCODE^12,13^. We focused on 855 unrelated European individuals with matching genotype and RNA-seq data (Methods). It is worthwhile noting that RNA-seq data was previously quantified using Gencode v19^19^ and RSEM^20^. We began with 24,905 genes and 93,293 isoforms in PsychENCODE. On average, each gene had four isoforms, but some genes had as many as 64 brain-expressed isoforms (**Figure S1**). Of these, 22,965 genes and 89,926 isoforms had converged *h*^2^_SNP_ estimates in the univariate model. Median *h*^2^_SNP_ estimates were 0.31 and 0.35 for gene and isoform expression, respectively (**Figure 2a**), and median *h*^2^_cis_ estimates were 0.01 and 0.01 for gene and isoform expression, respectively. 2,822 genes had significant *h*^2^_SNP_ at Bonferroni-adjusted *P* value < 0.05, while 3,557 genes had at least one isoform with *h*^2^_SNP_ significantly different from zero at Bonferroni-adjusted *P* value < 0.05. With a more lenient threshold at *P* < 0.05, 7,239 and 10,139 genes were heritable at the gene- and isoform-level, respectively. This increase in the number of heritable genes reflects added granularity with isoform-level analyses.

**Figure 2:**
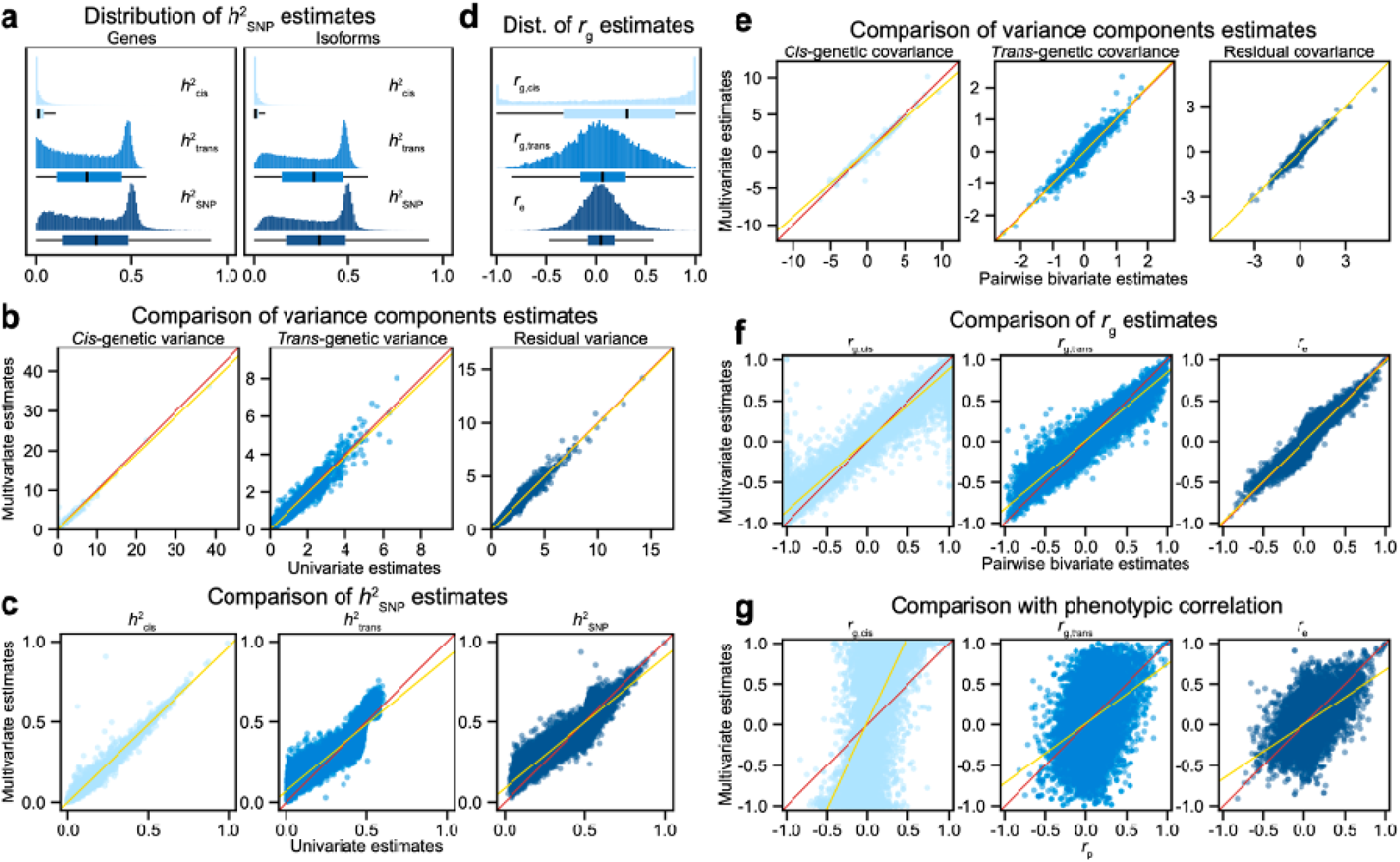
Comparison of *h*^2^_SNP_ and *r*_g_ estimates from different variance components models. **a**, Shown are the distributions of *h*^2^_SNP_ estimates for gene and isoform expression from fitting a univariate variance components model (22,965 genes + 89,926 isoforms). Note that *h*^2^_SNP_ = *h*^2^_cis_ + *h*^2^_trans_. **b**, Shown are variance components estimates from fitting univariate and multivariate variance components models. **c**, Shown are *h*^2^_SNP_ estimates from univariate and multivariate variance components models. **d**, Shown is the distribution of *r*_g_ estimates from fitting a multivariate variance components model. Note that *r*_g,cis_ captures aggregate degree of pleiotropy among *cis*-SNPs, while *r*_g,trans_ captures aggregate degree of pleiotropy among *trans*-SNPs. *r*_e_ captures residual correlation due to (unknown) biological and technological factors. The bimodal distribution of *r*_g,cis_ with two modes in the extremes suggests that there are substantial shared genetic influences among *cis*-SNPs. **e**, Shown are (co)variance components estimates from pairwise bivariate and multivariate variance components models. **f**, Shown are *r*_g_ estimates from pairwise bivariate and multivariate variance components models. **g**, Shown are *r*_g_ estimates from a multivariate variance components model and expression (phenotypic) correlation (*r*_p_). The same *r*_p_ values are plotted on the x-axis. All estimates are from REML estimation. The red lines are the diagonal lines, while the yellow lines denote the lines from linear regression.

It is standard practice when fitting linear mixed models to compare REML and ML estimates, since REML estimates tend to be less biased, while ML estimates can have lower mean squared error (MSE). For the univariate variance components model, we found strong concordance between REML and ML estimates (**Figure S2**). We note that *h*^2^_trans_ and *h*^2^_SNP_ estimates exhibit larger standard errors than *h*^2^_cis_ estimates (**Figure S2**), which is in line with expectation^21^, so we caution the readers in interpreting these *h*^2^_trans_ and *h*^2^_SNP_ estimates.

To ensure that our estimates were robust, we compared gene- and isoform-level *h*^2^_SNP_ estimates for genes with a single isoform (n = 7,246 genes). We found that they were highly concordant (**Figure S3**) despite gene- and isoform-level expression data being processed separately. When we fit a univariate variance components model with two variance components that only specifies *cis*-genetic effects, we observed concordant estimates for *h*^2^_cis_, although the estimates for the two variance components model were slightly inflated over the three variance components model, indicating the importance of correct model specification. Finally, we ran *cis*-eQTL analyses for gene- and isoform-level expression using QTLtools and found that most (if not all) heritable genes and isoforms harbor a significant eQTL (**Figure S4**).

Using QTLtools^18^, we identified 8,981 genes and 17,174 isoforms that harbor *cis*-eQTL at FDR < 0.05. The top associated SNP or index eQTL explained about 70% of variance from *h*^2^_cis_ (**Figure S5**). Conditional analyses using QTLtools found that most genes and isoforms have a single *cis*-eQTL signal, suggesting sparse and large genetic effects among *cis*-SNPs. We note that the number of independent *cis*-eQTL will likely increase with an increase in sample size, but in the present study, only 25% of genes and 15% of isoforms harbored more than one significant eQTL.

Next, we sought to investigate the polygenicity of *trans*-SNPs by partitioning genetic variances among 22 autosomal chromosomes. Since these estimates were noisy due to the limited sample size of PsychENCODE, we fit a penalized model with lasso penalty^22^ (Methods), which had an effect of shrinking variance components estimates and hence prioritizing the most important SNP effects. We found that the chromosome a given gene or isoform belongs to almost always had non-zero estimates, indicating substantial *cis*-SNP effects, while the rest of the chromosomes had non-zero estimates in proportion to their number of SNPs, suggesting polygenic effects (**Figure S6**). Altogether, *cis*-SNP effects were sparse but large, and *trans*-SNP effects were polygenic.

### Pleiotropy among brain isoform expression

Given their genomic proximity and co-regulation, we hypothesized that isoforms are under shared genetic influences. To address this question, for genes with multiple heritable isoforms at *P* < 0.05, we jointly modeled isoform expression using the multivariate variance components models, which allowed us to estimate *h*^2^_SNP_ and *r*_g_ together. Of note, 3,801 genes had at least two heritable isoforms, and 1,743 genes had at least three heritable isoforms at *P* < 0.05. Thus, our multivariate models included between two to as many as 23 isoforms. We observed concordant variance components (**Figure 2b**) and *h*^2^_SNP_ estimates (**Figure 2c**) between the univariate and multivariate models, although we note that *h*^2^_trans_ and *h*^2^_SNP_ estimates were slightly deflated for multivariate models compared to univariate models. The distribution of *r*_g,cis_ was bimodal with the two extremes (**Figure 2d**) and a median estimate of 0.31, suggesting that *cis*-SNPs tend to affect or co-regulate nearby isoforms together. The negative *r*_g,cis_ estimates close to -1 are characteristic of isoform switching events. Meanwhile, *r*_g,trans_ and *r*_e_ estimates followed unimodal distributions with their medians shifted right from zero (Figure 2d). This makes biological sense in that the distal genetic regulators and other biological factors tend to influence transcription of nearby isoforms together.

Next, we sought to understand if fitting pairwise bivariate or multivariate models lead to any meaningful differences in *r*_g_ estimates. This question is relevant for not only molecular readouts such as isoform expression, but also for complex traits and diseases in general, because most of the genetic correlation estimates are based on fitting pairwise bivariate models^3,5,6,23^. As for isoform expression, covariance terms in variance components estimates were concordant (**Figure 2e**), while *r*_g_ estimates were slightly inflated for pairwise bivariate models compared to multivariate models (**Figure 2f**). Further, we observed numerous cases where the signs of *r*_g_ estimates flipped between these two models. Based on these findings and previous observations that pairwise bivariate models tend to yield estimates that do not respect the positive semidefinite constraint of variance components parameters^8^, we construe that multivariate estimates are generally more accurate than pairwise bivariate estimates.

Previous work demonstrated that genetic correlation mimics phenotypic correlation in the direction of effect^24-30^, and thus motivating the use of phenotypic correlation as a proxy for genetic correlation. We sought to test this hypothesis for isoform expression and we observed that genetic correlation (both *r*_g,cis_ and *r*_g,trans_) generally recapitulates phenotypic correlation (or co-expression) in the direction of effect with *r*_g,cis_ being overall larger in magnitude than phenotypic correlation (**Figure 2g**). This is consistent with previous studies and our own observation that the isoforms of a given gene are under substantial shared genetic influences, particularly among *cis*-SNPs (**Figure 2d**).

### *ATP9B* gene as a case study for *h*^2^_SNP_ and *r*_g_ analyses

We now focus on *ATP9B* gene to contextualize our *h*^2^_SNP_ and *r*_g_ results (**Figure 3**). In Gencode v19, *ATP9B* had 27 annotated isoforms, 17 of which were determined to be brain-expressed in PsychENCODE based on the criteria TPM > 0.1 in at least 25% of samples^12^. Nine of these isoforms were found heritable at *P* < 0.05 after fitting the univariate variance components model (**Figure 3a**). Pairwise bivariate and multivariate models were fit for the nine heritable isoforms, which resulted in concordant *h*^2^_SNP_ estimates with similar magnitudes (**Figure 3b**). Standard errors for *h*^2^_SNP_ estimates were comparable across univariate, pairwise bivariate, and multivariate models, while standard errors for *h*^2^_trans_ were consistently larger than those of *h*^2^_cis_. Because we modeled nine isoforms, we note that there were eight pairwise bivariate estimates for each *h*^2^_SNP_ parameter (**Figure 3b**). For the *r*_g_ parameter, pairwise bivariate and multivariate models yielded estimates that were concordant in direction of effect, but less so in magnitude. That is, pairwise bivariate estimates were relatively inflated compared to multivariate estimates for both *r*_g,cis_ and *r*_g,trans_ (**Figure 3c**). Meanwhile, for both pairwise bivariate and multivariate models, *r*_g,cis_ estimates were substantially larger than *r*_g,trans_ estimates, suggesting that there were shared genetic influences, particularly among *cis*-SNPs. Not to mention, *r*_g,cis_ and *r*_g,trans_ estimates were generally larger than *r*_e_ estimates. Further comparison of *r*_g_ and *r*_e_ estimates with *r*_p_ estimates revealed that phenotypic correlation resembled residual correlation most closely (**Figure 3d**), indicating that for this particular gene *ATP9B*, residual (e.g. non-genetic biological and technological) factors overall exert larger effects than genetic factors (i.e. *cis*- and *trans*-SNPs). We note that one might be confused by this observation of high *r*_g,cis_ estimates but little resemblance between *r*_g,cis_ and *r*_p_ estimates. This is possible because *r*_g_ and *r*_e_ estimates are on a standardized scale, and hence the magnitude of residual variance-covariance can be larger than genetic variance-covariance while *r*_e_ is smaller than *r*_g_^30^. Finally, upon visual inspection of eQTL results, we found that there were distinct isoform-level eQTL signals (**Figure 3e**). For *ATP9B*, gene-level expression was also heritable and harbored strong eQTL signals (**Figure 3e**) unlike in subsequent examples.

**Figure 3:**
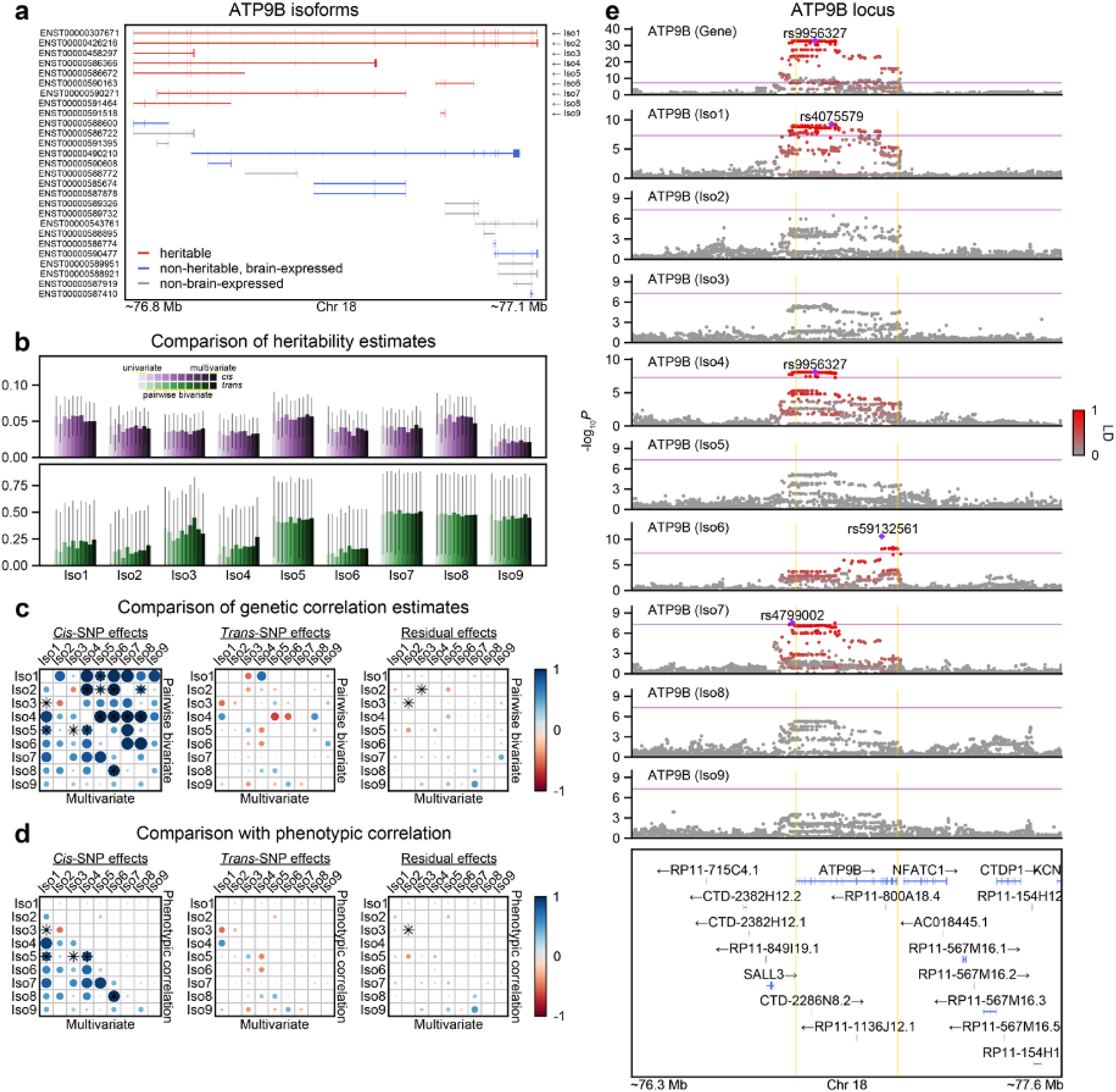
*ATP9B* as an example gene in *h*^2^_SNP_, *r*_g_, and *cis*-eQTL analyses. **a**, Shown are *ATP9B* isoforms that are present in Gencode v19. Nine isoforms that are found heritable in a univariate variance components model with a variation of the likelihood ratio test (LRT)^17^ are highlighted in red, and eight isoforms that are determined to be brain-expressed^12^ but not heritable are highlighted in blue. **b**, *h*^2^_cis_ and *h*_2trans_ estimates are shown in top and bottom rows, respectively. Colors encode estimates from univariate, pairwise bivariate, and multivariate variance components models. Because there are nine isoforms that are modeled jointly, there are eight pairwise bivariate estimates. All error bars denote ± one standard errors calculated from the Fisher information matrix. **c**, Shown are *r*_g,cis_, *r*_g,trans_, *r*_e_ estimates from pairwise bivariate and multivariate variance components models for *ATP9B*. The lower triangular elements represent multivariate estimates, while the upper triangular elements represent pairwise bivariate estimates. Asterisks denote significance from the Wald test at *P* < 0.05. **d**, Alike panel **c**, except that the upper triangular elements now represent phenotypic correlations (*r*_p_). The same *r*_p_ values are plotted. **e**, LocusZoom plot for *ATP9B* gene and isoform expression. Top row shows gene-level results. Index SNPs for features passing *P* = 5 × 10^−8^ threshold are shown and corresponding LD between other SNPs are displayed with the intensity of red color. Purple line denotes significance of *P* = 5 × 10^−8^, and yellow lines denote gene start and end sites for *ATP9B* gene. LD is calculated with individuals of European ancestry in the 1000 Genomes Project reference panel.

### Isoform-level eQTL signals prioritize candidate causal genes in GWAS loci

We next sought to understand whether isoform-resolution analyses could help prioritize disease genes in GWAS loci. Here, as a proof of concept, we nominate and share four such cases. The first is *XRN2* gene, which resides in one of two GWAS loci for autism spectrum disorder (ASD)^14^. Of note, the other ASD GWAS locus is a highly pleiotropic genomic region characterized by long-range, complex LD^31^ (**Figure S7**), which complicates fine-mapping and gene prioritization efforts. For *XRN2*, there were three brain-expressed isoforms, one of which was significantly heritable (ENST00000430571; *h*^2^_cis_ = 0.07, *h*^2^_trans_ = 0.46, *P* = 5.3 × 10^−13^). eQTL signal of this isoform colocalized^32^ strongly with the ASD GWAS signal (PP4 = 0.78; **Figure 4a**; **Figures S8-9**), suggesting that this isoform might be the causal isoform underlying the ASD GWAS signal. The second example is *SYNE1* gene, a SFARI ASD risk gene, of which five isoforms were heritable (**Figure S10**). Three of these isoforms harbored strong and distinct eQTL signals, one of which colocalized with a well-known bipolar disorder (BD) GWAS signal^33^ (PP4 = 0.90; **Figure 4b**; **Figures S11-12**). The third example is *TBL1XR1* gene, another SFARI gene, for which loss-of-function is implicated in increased risk for ASD^34^ and developmental delay disorder (DDD)^35^. Two *TBL1XR1* isoforms were found heritable, and they harbored distinct isoform-level eQTL signals, one of which colocalized with a SCZ GWAS signal^36^ (PP4 = 0.44**; Figure 4c**; **Figure S13-14**). The last example is *SYT1* gene, which is another SFARI gene and a high-confidence DDD risk gene^35^. Three *SYT1* isoforms were found heritable (**Figure S15**), two of which colocalized with an educational attainment (EA) GWAS signal^37^ (PP4 = 1.0; **Figure 4d**; **Figures S16-17**).

**Figure 4:**
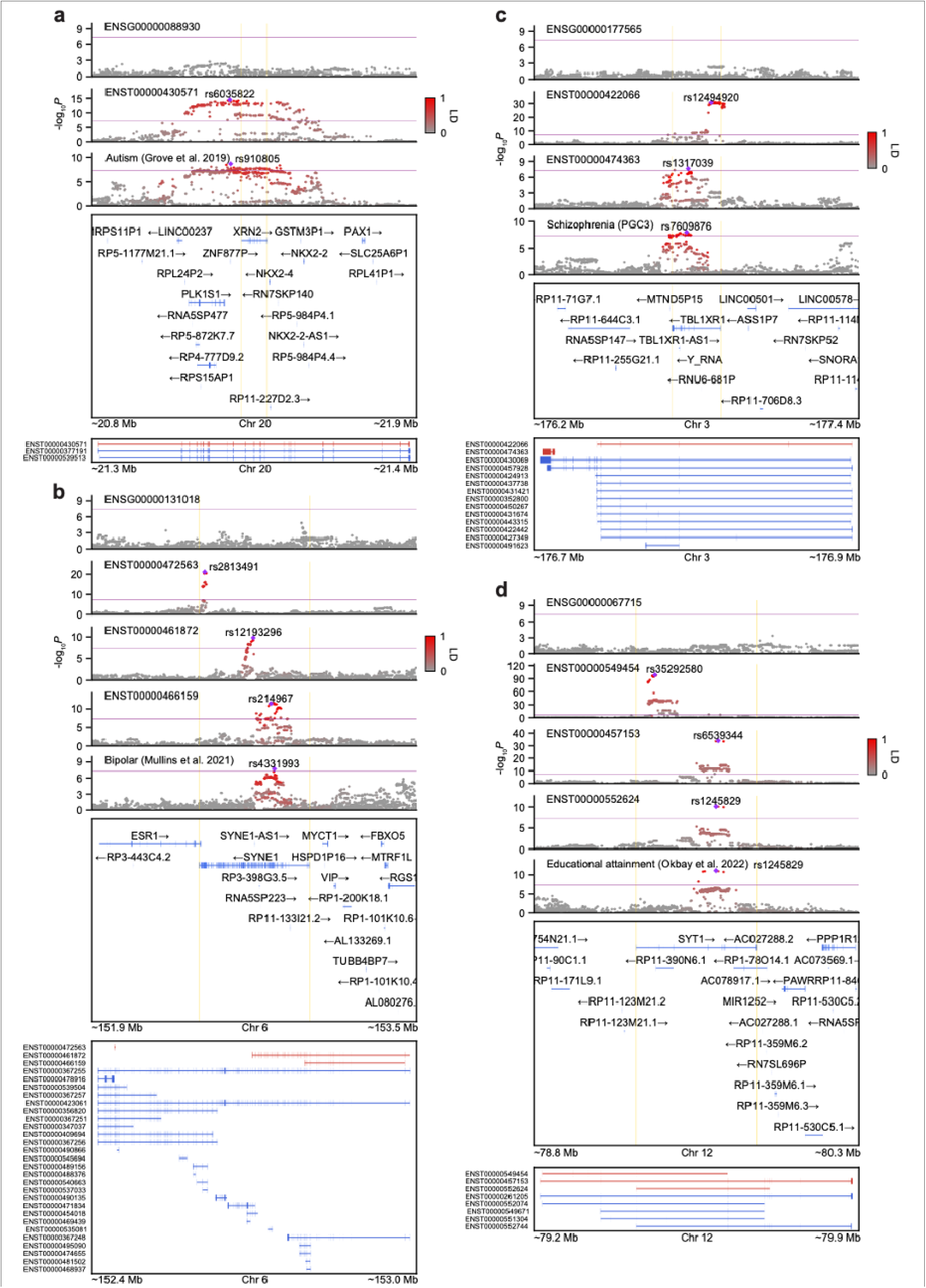
Isoform-level eQTL signals prioritize candidate causal genes in established GWAS loci. Shown are LocusZoom plots for **a**, *ATP9B*, **b**, *SYNE1*, **c**, *TBL1XR1*, and **d**, *SYT1* genes along with several complex phenotypes. Select isoforms that are included in LocusZoom plots are highlighted in red, while all other isoforms that are brain-expressed are highlighted in blue. Due to space constraints, isoforms that are present in Gencode v19 but not found brain-expressed are omitted. Index SNPs for features passing *P* = 5 × 10^−8^ threshold are shown and corresponding LD between other SNPs are displayed with the intensity of red color. Purple line denotes significance of *P* = 5 × 10^−8^, and yellow lines denote gene start and end sites. LD is calculated with individuals of European ancestry in the 1000 Genomes Project reference panel.

Given the strong implication of these example genes in neurodevelopmental disorders and their proximity to corresponding GWAS signals, these genes are likely the true causal genes driving the GWAS signals. Relatively straightforward patterns of linkage disequilibrium (LD) and reasonable gene densities also help in reaching this conclusion (**Figure 4**). It is important to note that all these example genes were missing gene-level eQTL signals, while harboring isoform-level eQTL signals, which highlights the potential of isoform-level analyses. Due to incomplete brain isoform annotations, the isoforms we identify may not be the true causal isoforms driving the GWAS signals, but with much more refined and complete transcriptome annotations along with more accurate quantifications of isoform-level expression (e.g. from long-read RNA-seq), we hypothesize that we would gain more fine-grained resolution in both gene- and isoform-level expression and thereby uncover more disease genes in GWAS loci.

### Replication of *XRN2* isoform-level eQTL signals in the developing human brain

As another proof of concept, we next sought to definitively fine-map the *XRN2* locus for ASD with improved isoform annotations. This was motivated by relatively simple gene structure of *XRN2*, for example, compared to that of *SYNE1* (**Figure 4b**). Further, accurate quantification of isoform expression in short-read RNA-seq data is challenging, given the presence of multi-mapped reads. The state-of-the-art isoform quantification methods^20,38,39^ address multi-mapped reads by probabilistically assigning them via maximizing the log-likelihood of the underlying statistical model with the EM algorithm. Unfortunately, this approach is valid only when all isoforms of a given transcriptome are known^40^. For example, when the transcriptome annotation is incomplete, multi-mapped reads that belong to an “unknown isoform” may be erroneously assigned to different isoforms that are annotated, thereby biasing these estimates. Accordingly, more complete transcriptome annotations are necessary to increase the accuracy of isoform quantification.

In Gencode v19, there were three isoforms for *XRN2*, all of which were determined to be brain-expressed. Upon close inspection of *XRN2* exons, we observed that the only difference between the ASD-associated isoform ENST00000430571 and the canonical isoform ENST00000377191 was skipped exon 2 (**Figure 4a**). On the contrary, the difference between the remaining isoform ENST00000539513 and the canonical isoform was a different exon 1 and the rest of the sequence was the same. Interestingly, we then noticed that all isoforms except the canonical isoform had been removed in subsequent Gencode versions past v19 due to low transcript support level. To ensure that our isoform is expressed and to improve the transcriptome annotation of the human brain, we combined the latest Gencode v40 annotation with existing long-read RNA-seq data from six studies^41-43^ (Methods). The data for these studies were mostly generated from fetal and adult human brain samples. To compile a list of high-confidence isoforms, we filtered for isoforms that were found in at least two or three different sources. Remarkably, we recovered all three *XRN2* isoforms in Gencode v19 with ENST00000377191, ENST00000430571, and ENST00000539513 found in six, four, and two studies, respectively (**Figure 5a**).

**Figure 5:**
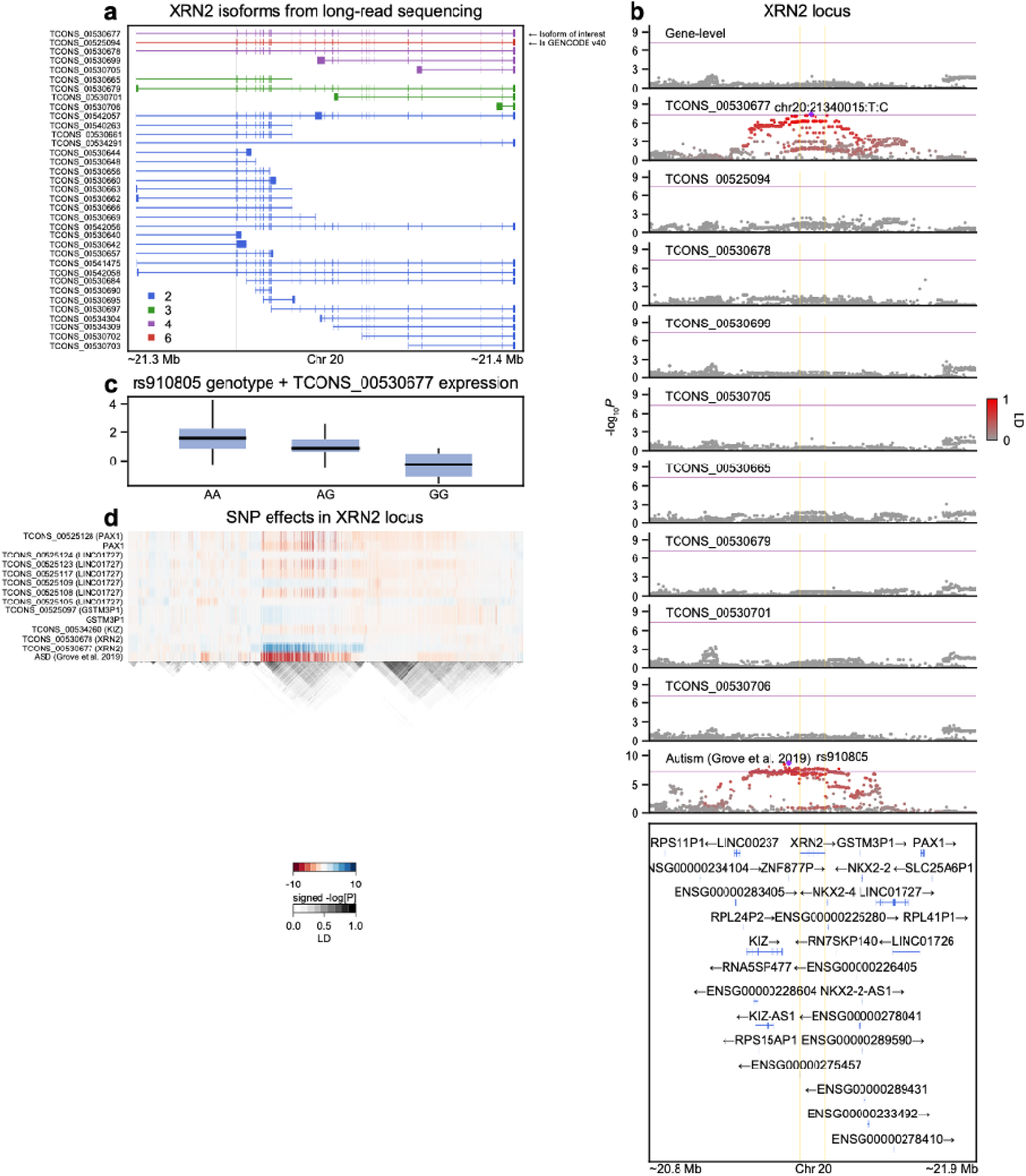
Fine-mapping of top ASD GWAS locus with isoform-level eQTL signals and improved isoform annotations in an independent fetal human brain dataset. **a**, Shown are high-confidence, credible *XRN2* isoforms compiled from six existing long-read studies^41-43^. Isoforms are colored with respect to the number of studies each isoform is found in. TCONS_00530677, TCONS_00525094, and TCONS_00541475 correspond to ENST00000430571, ENST00000377191, and ENST00000539513 in Gencode v19, respectively. The skipped exon 2 in TCONS_00530677 is shaded in grey. We subsequently filtered for isoforms found in at least three different studies and updated the transcriptome accordingly. **b**, Shown is LocusZoom plot for *XRN2* gene and its isoform expression based on the updated transcriptome annotation. Isoform expression was quantified in fetal human brain samples^47^. **c**, Index SNP for ASD GWAS (rs910805) and its risk increasing allele G was associated with reduced expression of TCONS_00530677. Note that G is also the minor allele. **d**, Shown is a heatmap of eQTL signals for both gene- and isoform-level expression for features within ±0.5 Mb window of (collapsed) gene start and end sites for *XRN2* gene. Although eQTL analyses were conducted for a total of 30 genes within this region and all their isoforms, we only plot the results of features with minimum *P* values less than 10^−4^ for visual clarity. For isoforms, their cognate genes are shown in parentheses. LD is calculated with 86 individuals of European ancestry in ref which are the same individuals used for eQTL analyses. These analyses were conducted using the GRCh38 human genome build, which leads to slightly different genomic coordinates relative to previous figures that were based on the hg19 reference genome.

Given that ASD genetic risk factors are known to converge in cell-types and biological pathways during brain development^44-46^, we then sought to replicate our adult brain isoform-level eQTL findings in fetal brain samples^47^. The previous work^47^ performed isoform-level eQTL analyses with Gencode v23 and did not detect any significant association for *XRN2*, presumably because ENST00000377191 is the only isoform present in that version of Gencode. In contrast, when we re-quantified fetal brain RNA-seq data, using Salmon^39^ with the updated transcriptome annotation, and repeated isoform-level *cis*-eQTL analyses, we observed a strong and specific eQTL signal for ENST00000430571 (or TCONS_00530677 equivalently) (**Figure 5b**). Further, the index SNP for ASD GWAS result (rs910805) and its risk increasing (minor) allele G was associated with reduced expression of ENST00000430571 (**Figure 5c**), which direction of effect is consistent with the adult brain findings (**Figure 4a**). As another sanity check, we tested all genes within ±0.5 Mb window of *XRN2* and their constituent isoforms from the updated transcriptome annotation and observed that no other gene- or isoform-level expression harbored eQTL signals that colocalized with the ASD GWAS signal (**Figure 5d**). Altogether, we replicated our adult brain *XRN2* findings with more complete isoform annotations in an independent fetal brain dataset and confidently narrowed down expression changes in ENST00000430571 as the causal signal for the ASD GWAS signal.

The *XRN2* isoform ENST00000430571 has exon 2 skipped, which in theory should be uncovered by annotation-free methods that detect local splicing patterns^48^. Indeed, the intron that corresponds to the end of exon 1 and the start of exon 3 were shown to harbor a significant splicing QTL (sQTL) in previous studies^45,49,50^. However, several other studies failed to detect a significant sQTL^2,51^. We suspect that this discrepancy is due to a combination of low level of expression for ENST00000430571 and differences in RNA-seq library preparation across studies with polyA selection methods being more susceptible to 3’ bias than rRNA depletion methods. Indeed, changes in splicing patterns between rs910805 genotypes were subtle (**Figure S18**), which could be overshadowed by polyA selection methods that induce 3’ bias.

## Discussion

In this study, we introduce a statistical method for fitting multivariate variance components linear mixed models that allows for arbitrary number of traits and variance components^10^. This method permits more flexible multi-trait analyses. We note that our method is not scalable to biobank-level data, but it is competitive on large data problems, and we recommend applying this method for datasets of size up to *n × d* = 50000, where *n* is the number of samples and *d* is the number of phenotypes. By leveraging this method and the functional genomic dataset from PsychENCODE^12,13^, we jointly model isoform expression—for the first time—and dissect the genetic influences (i.e. *h*^2^_SNP_ and *r*_g_) on human brain gene and isoform expression. We find a substantial proportion of the human brain transcriptome to be under genetic control. We find *cis*-SNP effects to be sparse and large individually, while *trans*-SNP effects to be polygenic and large in aggregate. We conclude that the isoforms of a given gene are under shared genetic influences, particularly among *cis*-SNPs. By comparing pairwise bivariate to multivariate models, we empirically show that multivariate models yield more accurate estimates. Finally, several genes are found heritable only at the isoform-level and their genetic signals colocalize strongly with GWAS signals, suggesting that isoform expression changes might be the underlying drivers of these GWAS signals. One notable example is *XRN2*, a specific isoform of which reduced expression appears to increase ASD risk. Overall, the present study comprehensively estimates genetic parameters for brain gene and isoform expression and highlights the utility of conducting isoform-resolution analyses.

We first note a major and important caveat of isoform-level analyses: the underlying generative model for the state-of-the-art short-read RNA-seq quantification tools^20,38-40^, start with an assumption that all expressed isoforms of a given transcriptome are known. But the human transcriptome annotation is far from complete, especially for the brain, which can result in inaccurate isoform quantifications and hence inaccurate downstream analyses. However, this also means that with more complete transcriptome annotations, we can obtain more accurate estimates of isoform-level expression even with short-read RNA-seq data^40^. Obviously, it is impossible to account for all possible (known and unknown) sample- and isoform-specific biases when resolving multi-mapped short-read RNA-seq reads and estimating isoform-level expression, but the current methods^20,38,39^ offer a reasonable starting point for running isoform-level analyses in conjunction with traditional gene-level analyses. Besides, isoforms are the fundamental biological units expressed in cells and in contrast to usual sQTL analyses^2,45,49-51^, isoform-level analyses are more interpretable in that they can pinpoint specific isoforms that are influenced by genetic variation. With extensive efforts to comprehensively catalog all brain-expressed isoforms^41-43,52,53^, we foresee that the accuracy of isoform-level quantification will improve for even short-read RNA-seq data.

The current study is limited by its sample size. This is evident by relatively large standard errors in our estimates, particularly for *trans*-SNP effects^21^. Hence with larger sample sizes, we expect to uncover many more genes and isoforms that are genetically regulated. We also note that the PsychENCODE dataset is an output of mega-analysis of six different studies, which can lead to decreased signal-to-noise ratio from heterogeneity and various technical artifacts. We envision increased genetic discovery with more homogenous genotype and RNA-seq data that have sufficient sample sizes. Moreover, the choice of RNA-seq normalization methods can have an impact on *h*^2^_SNP_ and *r*_g_ estimates. The PsychENCODE expression data was processed in log_2_-CPM-TMM scale, which better accounts for differences in library composition between samples in a large mega-analysis dataset such as PsychENCODE, but complicates the relationship between gene and isoform-level expression, including *h*^2^_SNP_ estimates (Methods). In the main text, we only highlight a handful of genes of which isoform expression changes seem to drive GWAS signals such as *XRN2, SYNE1, TBL1XR1*, and *SYT1*. Whether isoform-level genetic signals are more broadly enriched for disease risk and how many additional putative disease genes we can prioritize from isoform-resolution analyses remains to be further investigated.

Lastly, the *XRN2* gene encodes an RNA-binding protein that possesses 5’-3’ exoribonuclease activity. It is known to promote termination of transcription by degrading RNA to resolve R-loops. *XRN2* is mildly constrained (pLI = 0.35, LOEUF = 0.35), shows peak expression in the fetal brain^54^, and is enriched in neuronal cell-types^13,55^. Interestingly, visual inspection of phenome-scale LocusZoom plots reveals that there are two distinct GWAS signals within the *XRN2* locus, one for ASD and the other for blood cell-related phenotypes (**Figure S19**). Note that the same GWAS signal for ASD reached genome-wide significance in the latest GWAS for ADHD. We find that at the gene-level, *XRN2* is not heritable and does not harbor a significant eQTL, but at the isoform-level, one specific *XRN2* isoform is heritable and harbors a significant eQTL that colocalizes with the ASD GWAS signal. As a result of the skipping of the second exon, this isoform is missing 76 amino acids in the 5’ end compared to the canonical *XRN2* isoform. Structural and biological consequences of such changes in protein sequences need to be studied in the future. We note that the prioritized and canonical isoforms belong in different brain WGCNA modules (isoM23 and isoM35)^12^, which capture known neurobiological pathways and cell-types, potentially suggesting that the two isoforms might be involved in different biological pathways in the human brain.

## Data Availability

PsychENCODE genotype array and RNA-seq data are available at www.doi.org/10.7303/syn12080241 and processed summary-level data are available at Resource.PsychENCODE.org.

## Acknowledgments

This work was supported by the National Institute of Mental Health (T32MH073526 to M.K.; F30MH125523 to M.K.; R01MH121521 to M.J.G.; R01MH123922 to M.J.G.; P50HD103557 to M.J.G.) and the UCLA Medical Scientist Training Program (T32GM008042 to M.K.). Data were generated as part of the PsychENCODE Consortium, supported by: U01MH103392, U01MH103365, U01MH103346, U01MH103340, U01MH103339, R21MH109956, R21MH105881, R21MH105853, R21MH103877, R21MH102791, R01MH111721, R01MH110928, R01MH110927, R01MH110926, R01MH110921, R01MH110920, R01MH110905, R01MH109715, R01MH109677, R01MH105898, R01MH105898, R01MH094714, P50MH106934, U01MH116488, U01MH116487, U01MH116492, U01MH116489, U01MH116438, U01MH116441, U01MH116442, R01MH114911, R01MH114899, R01MH114901, R01MH117293, R01MH117291, R01MH117292 awarded to: Schahram Akbarian (Icahn School of Medicine at Mount Sinai), Gregory Crawford (Duke University), Stella Dracheva (Icahn School of Medicine at Mount Sinai), Peggy Farnham (University of Southern California), Mark Gerstein (Yale University), Daniel Geschwind (University of California, Los Angeles), Fernando Goes (Johns Hopkins University), Thomas Hyde (Lieber Institute for Brain Development), Andrew Jaffe (Lieber Institute for Brain Development), James Knowles (University of Southern California), Chunyu Liu (SUNY Upstate Medical University), Dalila Pinto (Icahn School of Medicine at Mount Sinai), Panos Roussos (Icahn School of Medicine at Mount Sinai), Stephan Sanders (University of California, San Francisco), Nenad Sestan (Yale University), Pamela Sklar (Icahn School of Medicine at Mount Sinai), Matthew State (University of California, San Francisco), Patrick Sullivan (University of North Carolina), Flora Vaccarino (Yale University), Daniel Weinberger (Lieber Institute for Brain Development), Sherman Weissman (Yale University), Kevin White (University of Chicago), Jeremy Willsey (University of California, San Francisco), and Peter Zandi (Johns Hopkins University). The Genotype-Tissue Expression (GTEx) Project was supported by the Common Fund of the Office of the Director of the National Institutes of Health, and by NCI, NHGRI, NHLBI, NIDA, NIMH, and NINDS.

## Author contributions

M.K. conceived the study. M.K. and H.Z. developed the software. M.K. and D.D.V. performed primary analyses. M.K. conducted data visualization. M.K. interpreted the results and wrote the manuscript with additional input from M.J.G. C.T.J., C.W., A.P. collected and managed long-read and fetal brain data. All authors read and approved the final manuscript.

## Methods

### The PsychENCODE genotype dataset

Genotype array and frontal cortex RNA-seq data from Freeze 1 and 2 of PsychENCODE were obtained from www.doi.org/10.7303/syn12080241. This consisted of uniformly processed data from six studies: BipSeq, LIBD_szControl, CMC-HBCC, CommonMind, BrainGVEX, and UCLA-ASD (see Table S1 and Fig. S33 in ref ^13^). Genotype data for these individual studies were previously harmonized through phasing and imputation with the Haplotype Reference Consortium (HRC) reference panel. We focused on 860 unique European individuals with matching genotype and frontal cortex RNA-seq data. We started with 5,312,508 HRC imputed SNPs and filtered for SNPs with minor allele frequency (MAF) > 0.01, genotype and individual missingness rate < 0.05, and Hardy-Weinberg equilibrium *P* values > 1e-6. Five pairs of individuals had classic genetic relationship matrix (GRM) values > 0.05 when using all filtered SNPs, while 647 pairs of individuals had GRM values > 0.025. We kept one individual from each of five pairs and only SNPs belonging to autosomal chromosomes, resulting in a total of 855 unrelated European individuals and 4,685,674 SNPs for downstream analyses.

### The PsychENCODE RNA-seq dataset

We used post-QC RNA-seq data that were fully processed, filtered, and normalized (see Materials/Methods and Fig. S3 in ref ^12^). Of note, RNA-seq reads were previously aligned to the hg19 reference genome with STAR 2.4.2a and gene and isoform-level quantifications calculated using RSEM v1.2.29. Genes and isoforms were filtered to include those with TPM > 0.1 in at least 25% of samples. Gene and isoform expression were separately normalized using TMM normalization in edgeR and log_2_-transformed. The same set of known biological and technical covariates were used for mean or fixed effects, which include age, age^2^, study, sex, diagnosis, RNA integrity number (RIN), RIN^2^, post-mortem interval (PMI), 24 sequencing principal components (PCs), and 5 genetic PCs. RNA-seq data was also restricted to frontal cortex samples from European individuals as well as genes and isoforms belonging to autosomal chromosomes, resulting in a total of 24,905 genes and 93,293 isoforms based on Gencode v19 annotations. The same expression data was used for all downstream analyses unless otherwise stated. We note that we did not correct for latent factors for the main analyses such as hidden covariates with prior (HCP)^56^, given that we are modeling *cis*- and *trans*-SNP effects simultaneously and we risk removing trans effects by adjusting for such latent factors^57^.

### Multiple variance components linear mixed model for univariate response

Given an *n* × 1 response vector **y** and predictor *n* × *p* matrix **X**, the variance components linear mixed model assumes ***y*** ∼ *N*(**Xβ, Ω**), where

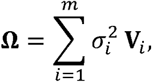

and **V**_1_, …, **V**_*m*_ are *m* known positive semidefinite matrices. The parameters of the model include mean effects (**β**) and variance components 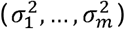. **Ω** is assumed to be positive definite. The simplest heritability and GWAS model assumes two variance components, where **V**_1_ is a kinship matrix and **V**_2_ = **I**. This model is misspecified for gene expression, since SNPs in the vicinity of a gene (i.e. *cis*-SNPs) are known to exert stronger effects on its expression. Therefore, for gene and isoform expression, we specified three variance components, where

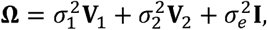

and **V**_1_ and **V**_2_ are empirical kinship matrices constructed from *cis*- and all other SNPs (i.e. *trans*-SNPs), respectively, such that 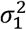 and 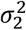 capture aggregate genetic effects of corresponding sets of SNPs. The classic genetic relationship matrix (GRM) was used for both empirical kinship matrices, under which 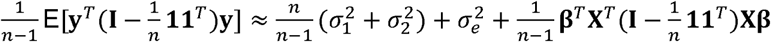. This equality is approximate, since the diagonal elements of GRM are not exactly one. When there are no mean effects or there is only the intercept term, the overall variance can be decomposed into the sum of variance components as above, so it makes natural sense to define SNP-based heritability as

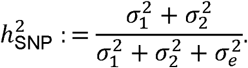

### Multiple variance components linear mixed model for multivariate response

The multivariate response variance components model assumes an *n* × *d* response matrix **Y** with vec **Y** ∼ *N(*vec(**XB**), **Ω**), where

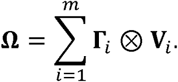

vec **Y** creates an *nd* × 1 vector from **Y** by stacking its columns and ⊗ denotes the Kronecker product. The parameters of the model include *p* × *d* mean effects **B** and *d* × *d* variance components that are (**Γ**_1_, …, **Γ**_*m*_) that are positive semidefinite. **Ω** is assumed to be positive definite. The univariate response model is subsumed under the multivariate response model when there is only a single phenotype. The simplest genetic correlation model looks at phenotypes pairwise and assumes two variance components. However, this model is likely inadequate for isoform-level expression, so for constituent isoforms of a given gene, we modeled them jointly and specified three variance components such that

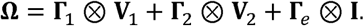

Corresponding to the univariate response model, **Γ**_1_ captures genetic variances and covariances among *cis*-SNPs, while **Γ**_2_ captures genetic variances and covariances among *trans*-SNPs. Let (**Y**)._*j*_ denote the *j*th column of **Y** and (**Y**)_*jk*_ the (*j,k*) th element of **Y**. Then 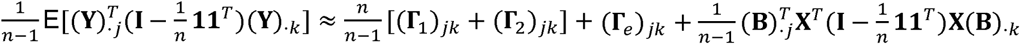. In other words, just as the overall variance could be decomposed into the sum of genetic variances and residual variance, the overall covariance can be decomposed into the sum of genetic covariances and residual covariance following this model. Genetic correlation is defined as genetic covariance divided by the product of square root of corresponding genetic variances. For example, genetic correlation for *j,k* th isoforms among *cis*-SNPs is 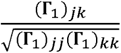. It is worthwhile to note that the magnitude of genetic covariance or genetic correlation can differ between *cis*- and *trans*-SNPs.

Residual variance could be further decomposed into **Γ**_*e*_= **Γ**_*τ*_ + **Γ**_*ϵ*_ where **Γ**_*τ*_ captures covariance from sampling error and measurement error from shared transcript structures among isoforms, while **Γ**_*ϵ*_ captures covariance arising from other biological factors. In practice, these two are indistinguishable from one another, but one could make use of bootstrap samples (i.e. also known as technical replicates) from Kallisto or Salmon to estimate. Then we can let 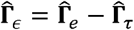.

### Relationship between gene and isoform expression 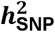

Gene-level expression in unit of transcripts per million (TPM) is conventionally assumed to be the sum of expression of its constituent isoforms in TPM. Suppose there are *d* isoforms for a given gene, then

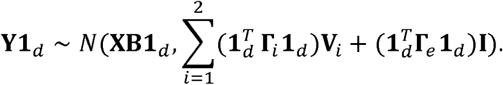

As a result, the overall variance in gene expression is approximately equal to 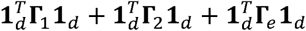 and gene-level SNP-based heritability becomes

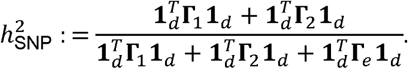

Four scenarios are possible based on the above equation: 1. both gene and isoforms are not heritable, 2. both gene and (a subset of) isoforms are heritable, 3. only gene is heritable, 4. only (a subset of) isoforms are heritable.

In practice, gene and isoform-level expression are log_2_ -transformed, which leads to the distortion of the above-mentioned relationship, where 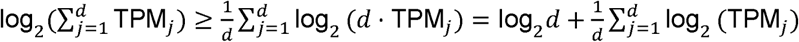. The PsychENCODE expression data was further processed not in TPM but instead in log -CPM-TMM, which normalization better accounts for differences in library composition across samples in a large mega-analysis dataset such as PsychENCODE, but complicates the relationship between gene and isoform-level expression.

### Estimation in variance components model

We consider both maximum likelihood (ML) and restricted (or residual) maximum likelihood (REML) approaches. REML estimates tend to give less biased estimates of variance components, while ML estimates can still have smaller mean squared error (MSE) and are useful for the likelihood ratio test (LRT). For the univariate response model, the log-likelihood function is

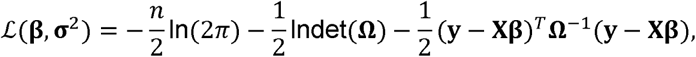

where 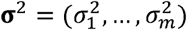. The corresponding score (gradient) vector is

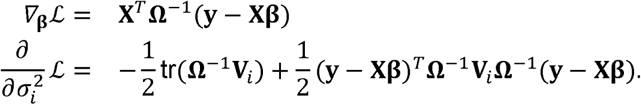

The observed information matrix has elements

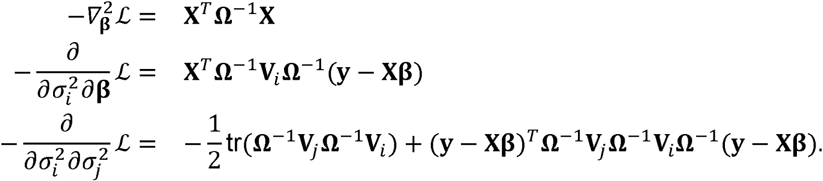

The expected (Fisher) information matrix has elements

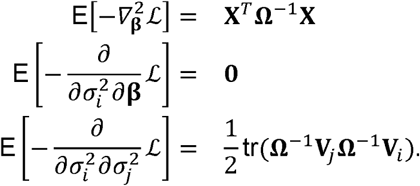

Suppose rank (**X**) = *r* and columns of **A**∈ ℝ ^*n* × (*n*−*r*)^ span the null space of **X**^*T*^, then the log-likelihood function for REML estimation is

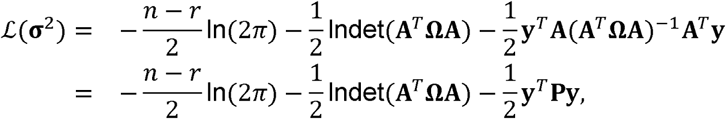

where **P** = **Ω**^−1^ − **Ω**^−1^**X**(**X**^*T*^ **Ω**^−1^ **X**)^−^**X**^*T*^ **Ω**^−1^. Due to the uniqueness of **P**, we can choose the columns of **A** to form an orthogonal basis such that **A**^*T*^**A** = 1. The score vector, observed and Fisher information matrices for this residual log-likelihood are similar to the above with **Ω** replaced by **A**^*T*^ **ΩA, V**_***i***_, by **A**^*T*^**V**_*i*_**A** for *i* = 1, …, *m* and **y** by **A**^*T*^**y**, and hence

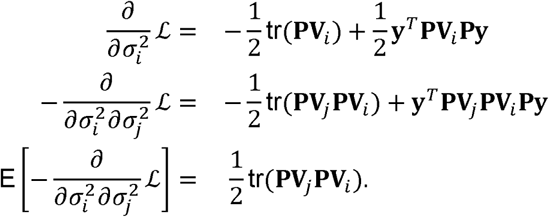

For the multivariate response model, the log-likelihood function is

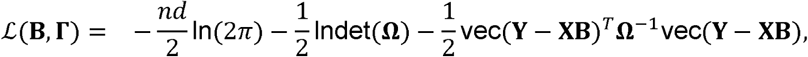

where **Γ** = (**Γ**_1_, …, **Γ**_*m*_) To ensure that **Γ**_*i*_ is positive semidefinite, we reparametrize with its Cholesky factor **L**_*i*_ such that 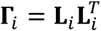 for *i* = 1, …, *m*. Then the score vector is

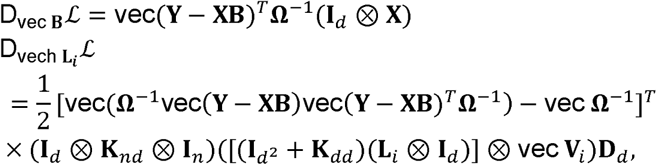

where **K**_*nd*_ is the *nd* × *nd* commutation matrix and **D**_*d*_ the *d*^2^ × *d* (*d* + 1)/2 duplication matrix.

Let 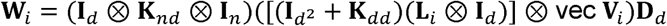 and **R** be the *n* × *d* matrix satisfying vec **R** = **Ω**^−1^vec(**Y** − **XB**). Then the observed information matrix has elements

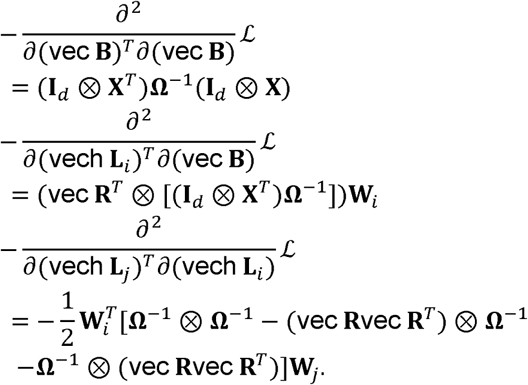

The expected (Fisher) information matrix has elements

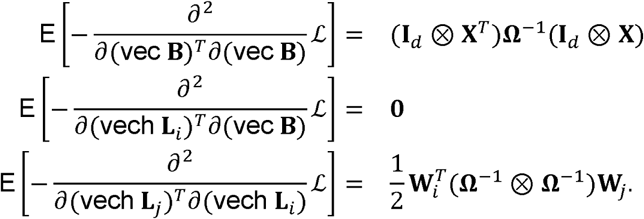

As a sanity check, one can observe that the above Fisher information boils down to the Fisher information for the univariate response model when *d* = 1. For REML estimation in the multivariate response model, the log-likelihood function is

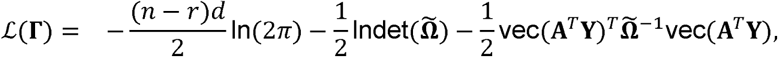

where 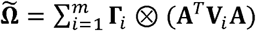. Let 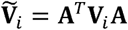 and 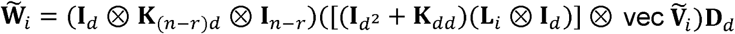, then the corresponding score vector, observed and Fisher information matrices are

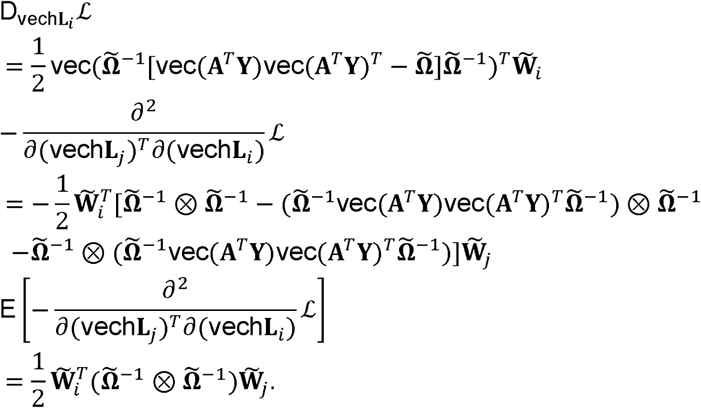

Variance components parameters are usually estimated using an iterative algorithm such as Fisher scoring and expectation-maximization (EM) algorithms. Fisher scoring uses the expected information matrix derived above instead of the observed information matrix in Newton’s method. To derive the EM algorithm, we let **Ω**_*i*_ = **Γ**_*i*_ ⊗ **V**_*i*_, *r*_*i*_ = rank(**V**_*i*_), *s*_*i*_ = rank(**Γ**_*i*_), and det^+^ (**Γ**_*i*_) denote the pseudo-determinant of **Γ**_*i*_ and 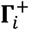 the pseudo-inverse of **Γ**_*i*_. Additionally, if we let vec **R**^(*t*)^ = **Ω**^−**(*t*)**^ vec(**Y** −**XB**^(*t*)^), then *Q*-function for the EM algorithm in the multivariate response model is

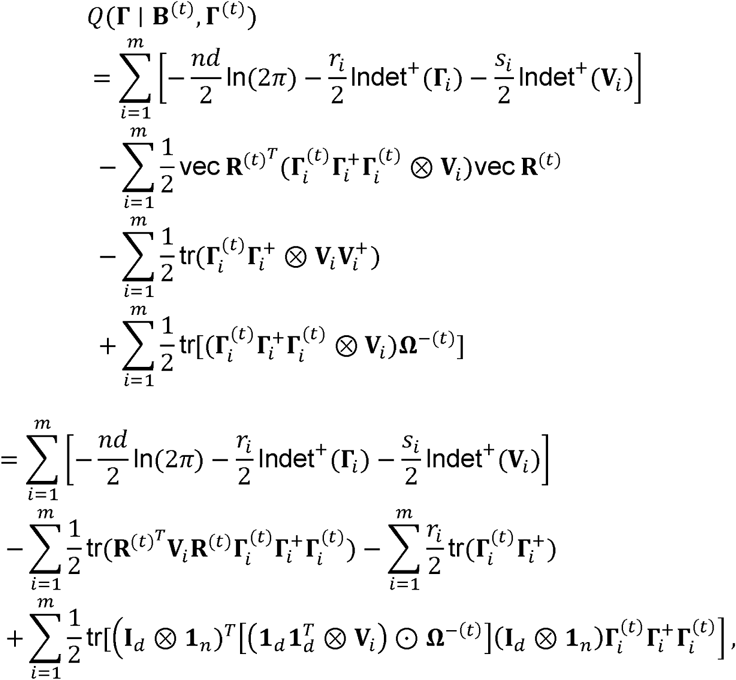

where ⊙ denotes the Hadamard (elementwise) product. Hence, the ECM updates are

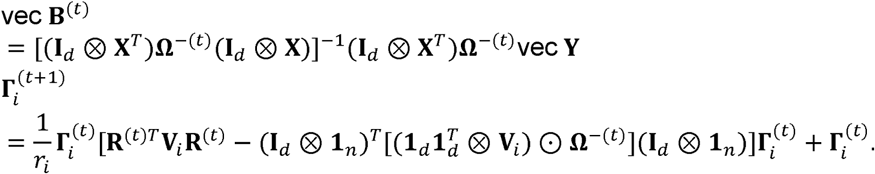

The EM algorithm is a special case of the minorization-maximization (MM) algorithm^9^. A different formulation of the MM algorithm^10^ implements a block ascent strategy by alternatively updating **β** and **σ**^2^ such that

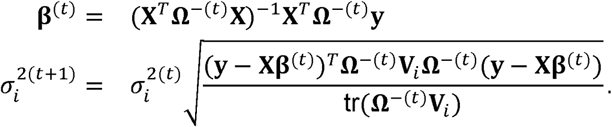

For the multivariate response model, the updates are

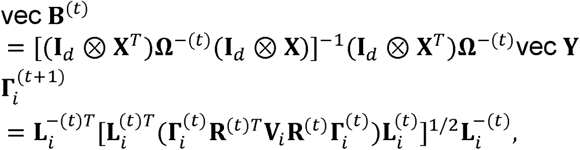

where 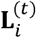 is the Cholesky factor of 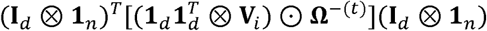 and vec **R**^(*t*)^ = **Ω**^−(*t*)^ vec (**Y** − **XB**^(*t*)^). As another sanity check, one can see that the above updates become the updates for the univariate response model when *d* = 1. For complete derivation, please see ref ^10^ and for REML estimation, the updates are basically of the same form. Compared to Newton’s method or Fisher scoring, the MM algorithm is numerically stable and computationally efficient. Although the number of iterations it takes is usually larger than Fisher scoring, the computational burden is smaller in each iteration^10^. The MM algorithm also gracefully respects the nonnegativity (or positive semidefiniteness) constraint of variance components. Compared to the EM algorithm, the MM updates converge more quickly, since the surrogate function hugs the log-likelihood function more tightly^10^. In the simplest heritability and GWAS model where there are two variance components, repeated matrix inversion can be avoided by the generalized eigenvalue decomposition of the two kernel matrices^10^. When there are more than two variance components, a matrix inversion is inevitable in each update, so the MM algorithm is not scalable to biobank-level data, and we recommend applying this method for datasets of size up to *n* × *d* = 50000. In this study, we take advantage of the aforementioned MM algorithm in estimating parameters in variance components models.

Univariate variance components linear mixed models were fit for gene and isoform-level expression with the above-mentioned mean effects covariates. We specified three variance components, two of which capture *cis*- and *trans*-SNP genetic effects. We defined *cis*-SNPs as those within 1 Mb window of gene start and gene end sites, and *trans*-SNPs as all the other SNPs. Based on this definition, 24,754 genes and 93,030 isoforms had non-zero *cis*-SNPs. The same set of *cis*-SNPs was used for a given gene and its constituent isoforms for direct comparison. The mean number of *cis*-SNPs were 3,264 and 3,274 for these genes and isoforms, respectively. Variance components parameters were estimated using both ML and REML. The maximum number of iterations was set to 3,000. In total, 22,965 genes and 89,926 isoforms had converged estimates. In most (if not all) cases where the estimates failed to converge, ML estimation was the issue.

As part of sensitivity analyses, we fit additional univariate models, including three variance components models with different definitions of *cis*-SNP windows (i.e. ±250 kb window and the entire chromosome), two variance components model with a single parameter capturing the entire genetic effects, and two variance components model with only *cis*-SNP effects (1 Mb window). In theory, model selection that best fits the data using a statistical criterion such as AIC metric is possible, but we did not conduct such analyses.

Multivariate variance components linear mixed models were fit for isoform-level expression. We specified three variance components, one of which captures *cis*-SNP genetic effects and the other *trans* effects. We used the same set of *cis*-SNPs that were used for univariate models. To reduce computational burden and the number of variance components parameters that need to be estimated, given limited sample size of the PsychENCODE dataset, we ran the multivariate model for isoforms with significant heritability estimates in a univariate model at *P* < 0.05. For isoforms that are perfectly correlated in expression, we included only one isoform of the two. Variance components parameters were estimated using REML. Note that the MM algorithm is numerically stable to fit even the non-heritable isoforms, but we chose not to. Lastly, pairwise bivariate variance components models were fit with the same scheme as multivariate models. For each gene with at least two heritable isoforms, the model was fit to all pairwise combinations of isoforms.

### Inference in variance components model

Inference on variance components parameters was done using a variation of the likelihood ratio test (LRT). Here, we tested the null hypothesis that both variance components corresponding to *cis*- and *trans*-SNP genetic effects are zero by fitting a null model with only a single residual variance components parameter. Then we compared the log-likelihood from ML estimation, which difference times two is assumed to follow a mixture of *χ*^2^ distributions^17^ as

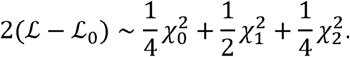

Standard errors for both genetic variances and genetic covariances were estimated using the Fisher information matrix, where

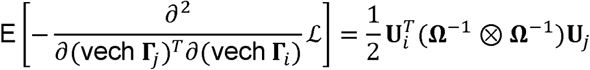

and 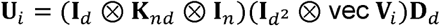. Note that this form is slightly different from the one shown earlier with **U**_*i*_ replaced by **W**_*i*_. This is because we previously imposed the positive semidefiniteness constraint by reparametrizing 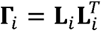. Then by using the vec-transpose operator,

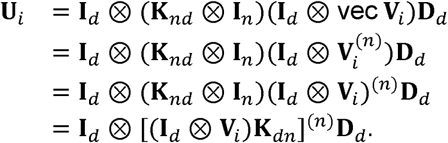

Naïve computation of the Fisher information matrix is not computationally feasible for even reasonable *nd*, so we must take advantage of the structure inherent in **U**_*i*_. Standard errors for heritability and genetic correlation estimates were subsequently calculated using the delta method. Inference on genetic covariance or genetic correlation parameters was done using the Wald test for both bivariate and multivariate models.

### Penalization of variance components

To investigate the polygenicity of gene and isoform expression, we also fit a univariate, 23 variance components model with separate SNP effects from each autosome such that

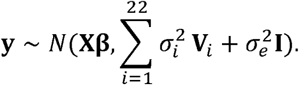

However, due to the limited sample size of PsychENCODE, these estimates were noisy and hence we instead minimized the lasso-penalized negative log-likelihood function^22^. One of the advantages of the MM algorithm is that it separates the parameters of a problem such that penalized estimation is conducive^10,22^. For each chromosome, we focused on heritable genes and isoforms (*P* < 0.05) that reside in that chromosome, and for these features, we calculated the number of times each chromosome appeared in the solution path.

### *cis*-eQTL analyses

We ran eQTL analyses with the same *cis*-SNPs used in variance components models for each gene and isoform using a linear model as implemented in QTLtools^18^. To account for linkage disequilibrium (LD), sample labels were permuted 1,000 times, and each iteration the most significantly associated *P* value was saved. Such a null distribution of *P* values was then fit to a beta distribution and the observed *P* value was subsequently adjusted to give an empirical *P* value. To account for multiple testing, empirical *P* values transcriptome-wide were FDR corrected. Conditional analyses were further performed through QTLtools to discover independently associated eQTL signals for gene and isoform expression. The top associated eQTL SNP for each gene and isoform was then used to calculate *R*^2^ or variance explained by index SNPs.

### Colocalization

We tested for colocalization between isoform-level eQTL signals and complex traits GWAS signals at the *XRN2, SYNE1, TBL1XR1*, and *SYT1* loci by using association results for SNPs within ±1 Mb window of respective gene bodies and present in both eQTL and GWAS results. We used the coloc.abf function in R package *coloc* v5.1.0.1^32^. This method assumes a single causal variant within each locus and assesses evidence of a shared association signal using a Bayesian approach. Sample size and MAF were used instead of sdY for *cis*-eQTL data and the default priors of *p*1 = *p*2 = 10^−4^ and *p*12 = 10^−5^ were applied.

### *XRN2* isoform re-quantification with long-read transcriptome annotation in fetal brain short-read bulk RNA-seq data

To better complete the human brain transcriptome annotation, we compiled long-read sequencing data of the human brain from six different studies^41-43^. This included in-house fetal brain single-cell Iso-Seq data and two adult human brain data shared by PacBio (https://www.pacb.com/connect/datasets/). Note that ref ^43^ includes data from non-brain tissues, while other studies are strictly from the adult or developing human brain. We started with finalized (e.g. filtered and merged) GTF files from each study, harmonized genomic coordinates with liftover to the GRCh38 human genome build, ran gffcompare v0.12.6 with Gencode v40 as a reference, and filtered for isoforms found in at least two or three studies. gffcompare searches for identical matches for internal exons, while it implements fuzzy matches for terminal exons, which is appropriate here since these exons can be sequenced to different lengths. We subsequently used the above two filtered/munged GTF files to re-quantify ref ^47^ fetal brain bulk RNA-seq data using Salmon v1.8.0. Gene-level expression was calculated to be the sum of its constituent isoform TPM. To repeat gene-level and isoform-level eQTL analyses, genotype data for ref ^47^ was filtered for typed variants with individual call rate (> 0.95), minor allele frequency (> 0.01), Hardy-Weinberg equilibrium (HWE) *P* value (> 10^−6^) and individuals based on genotype call rate (> 0.9) using PLINK 1.9. Genotype data was then phased and imputed with the TOPMed freeze 5 reference panel on the Michigan Imputation Server. After removing SNPs with low imputation quality (R2 < 0.3) and subsetting to 86 individuals of European ancestry, we again filtered SNPs based on individual call rate (> 0.95), MAF (> 0.05), HWE *P* value (> 10^−6^) and individuals based on genotype call rate (> 0.95). For eQTL analyses, we modeled either TPM or log_2_ TPM with age, sex, 4 genetic PCs, RIN, and 21 sequencing PCs from sequencing metrics generated from PicardTools as covariates.

## Data availability

PsychENCODE raw data are available at www.doi.org/10.7303/syn12080241 and processed summary-level data are available at Resource.PsychENCODE.org.

## Code availability

The Julia implementation of the MM algorithm for fitting multivariate variance components linear mixed models is available at https://github.com/Hua-Zhou/MultiResponseVarianceComponentModels.jl. The code used to perform bioinformatic analyses are available at https://github.com/mmkim1210/isoform-genetics. All plots were generated programmatically using Makie.jl^58^ and GeneticsMakie.jl^59^.

## Supplementary Figures

**Figure S1:**
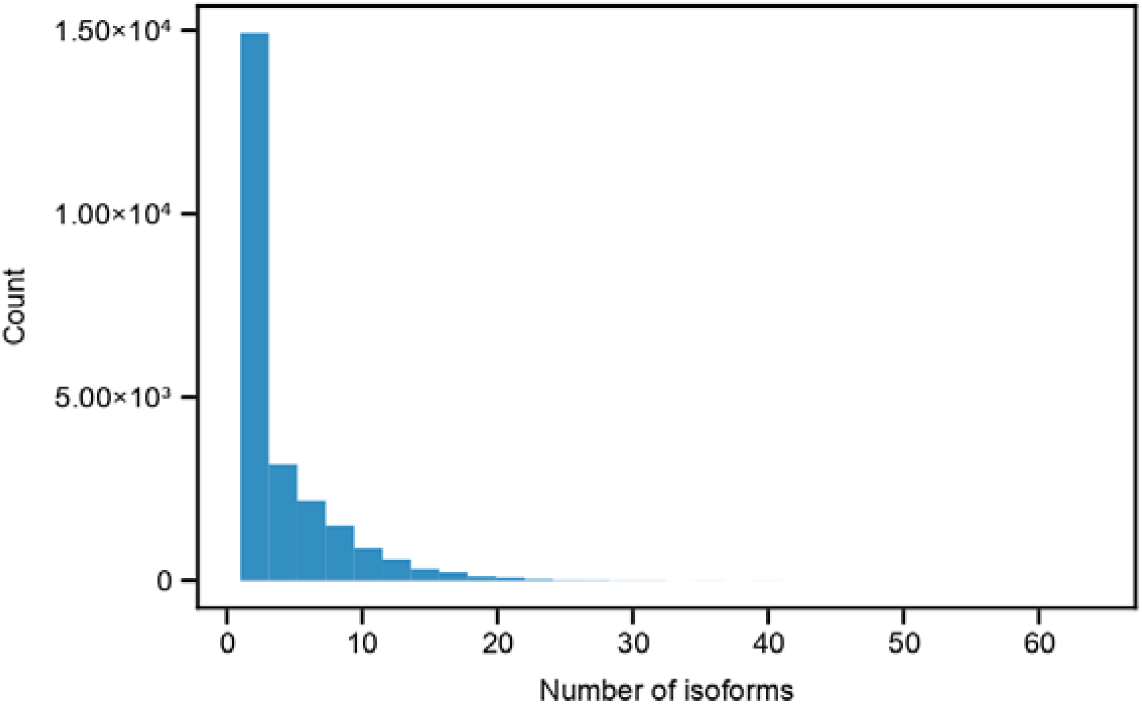
Distribution of the number of isoforms for 24,905 genes in PsychENCODE. The maximum number of isoforms is 64.

**Figure S2:**
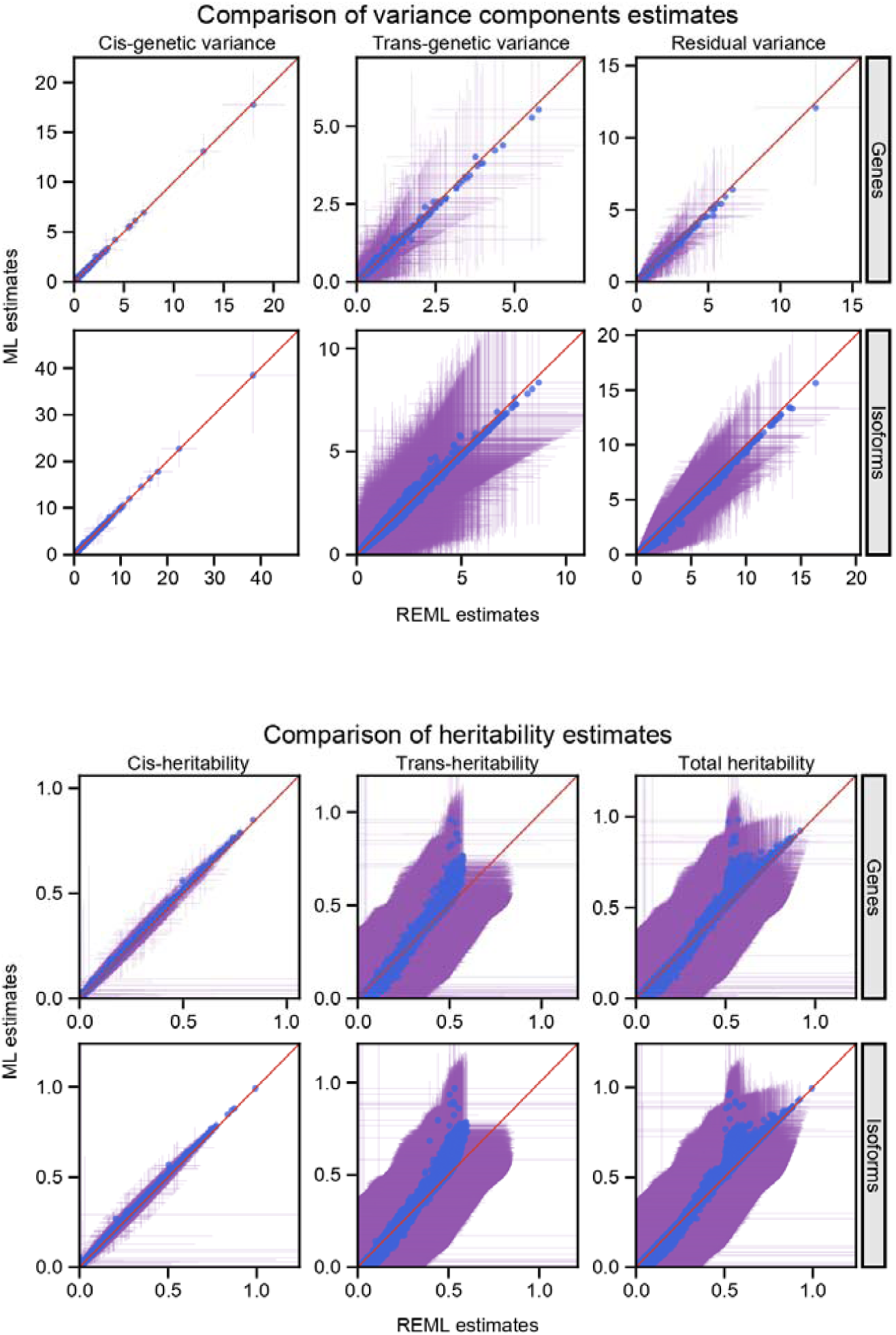
Comparison of REML and ML estimates in the univariate variance components models. REML and ML estimates were highly concordant for *h*^2^_cis_ and to a lesser degree for *h*^2^_trans_ and *h*^2^_SNP_. Error bars denote ± one standard errors. Note higher standard errors for *h*^2^_trans_ and *h*^2^_SNP_ relative to *h*^2^_cis_. Standard errors are calculated using the Fisher information matrix and the Delta method.

**Figure S3:**
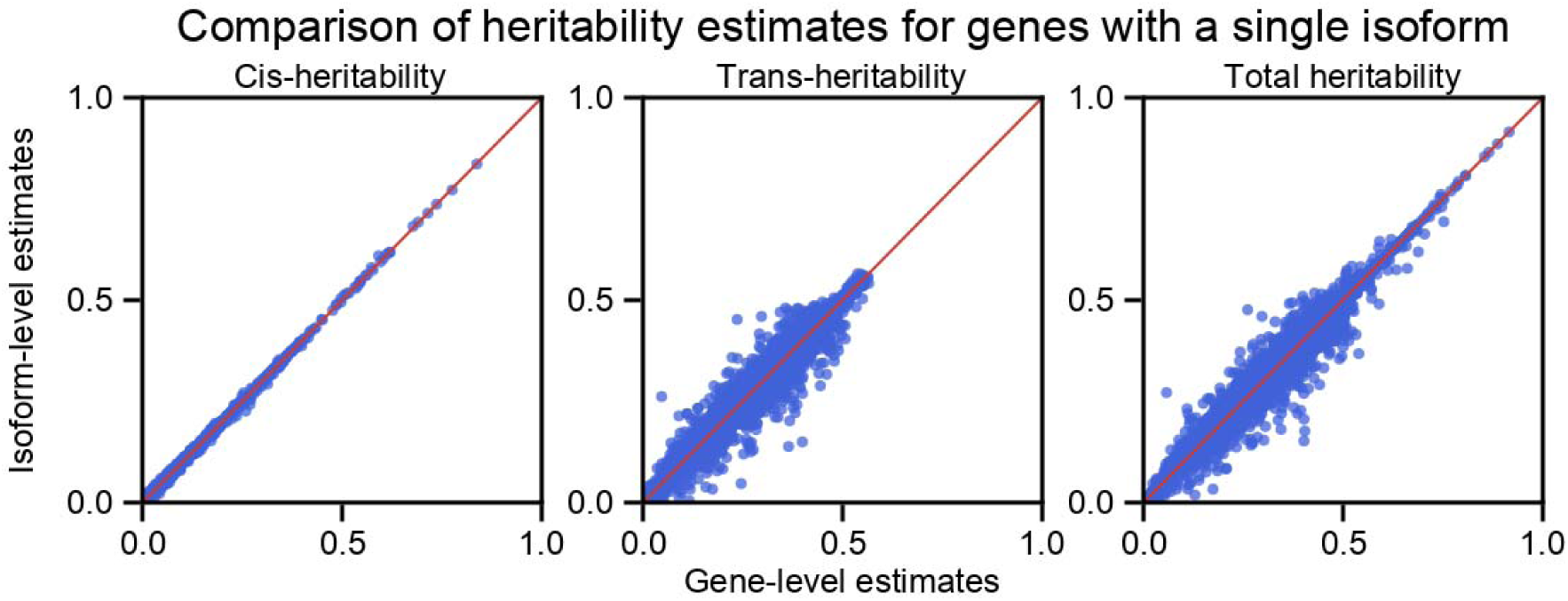
Comparison of heritability estimates for genes with a single isoform. *h*^2^_cis_ estimates were highly concordant, while more variability was observed for *h*^2^_trans_ and *h*^2^_SNP_ estimates.

**Figure S4:**
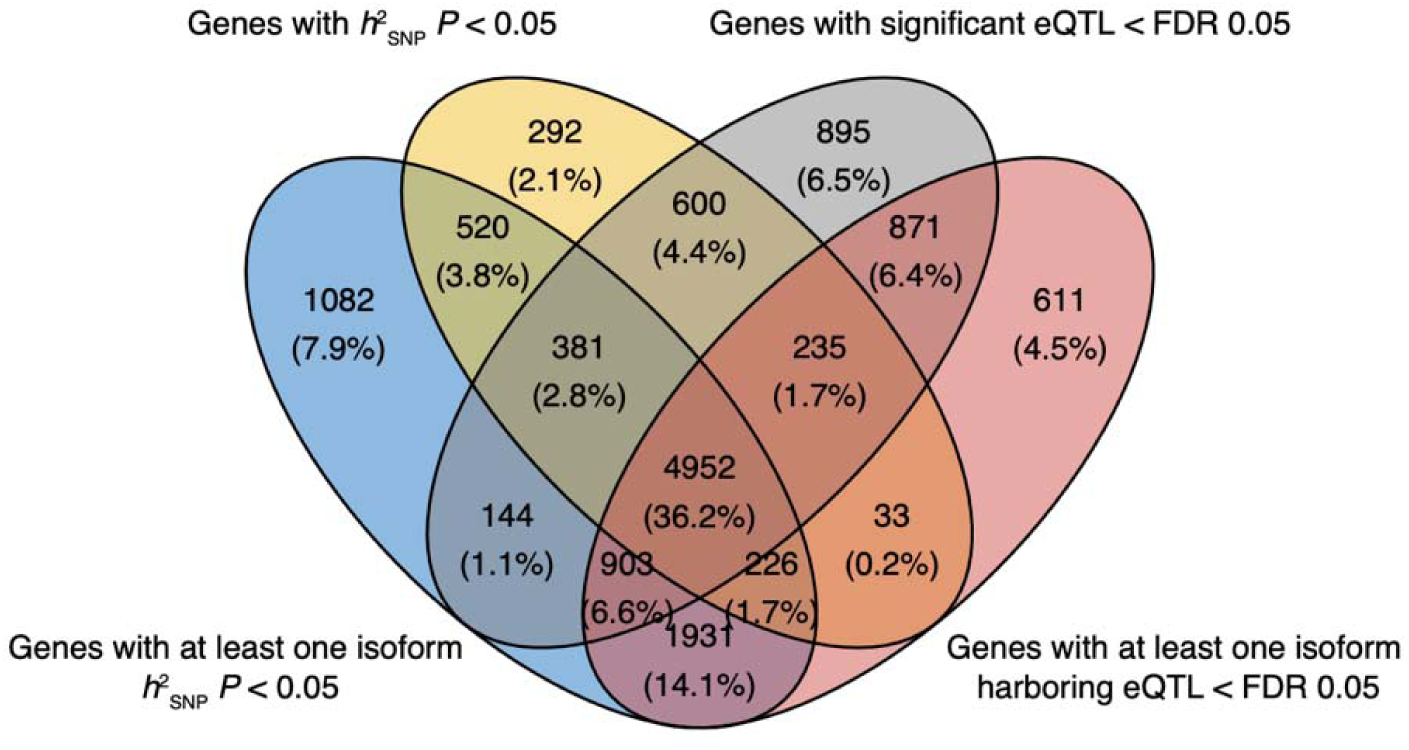
Comparison between heritability and *cis*-eQTL results. Shown is Venn diagram of overlap between heritable **g**enes and genes harboring eQTL at FDR < 0.05.

**Figure S5:**
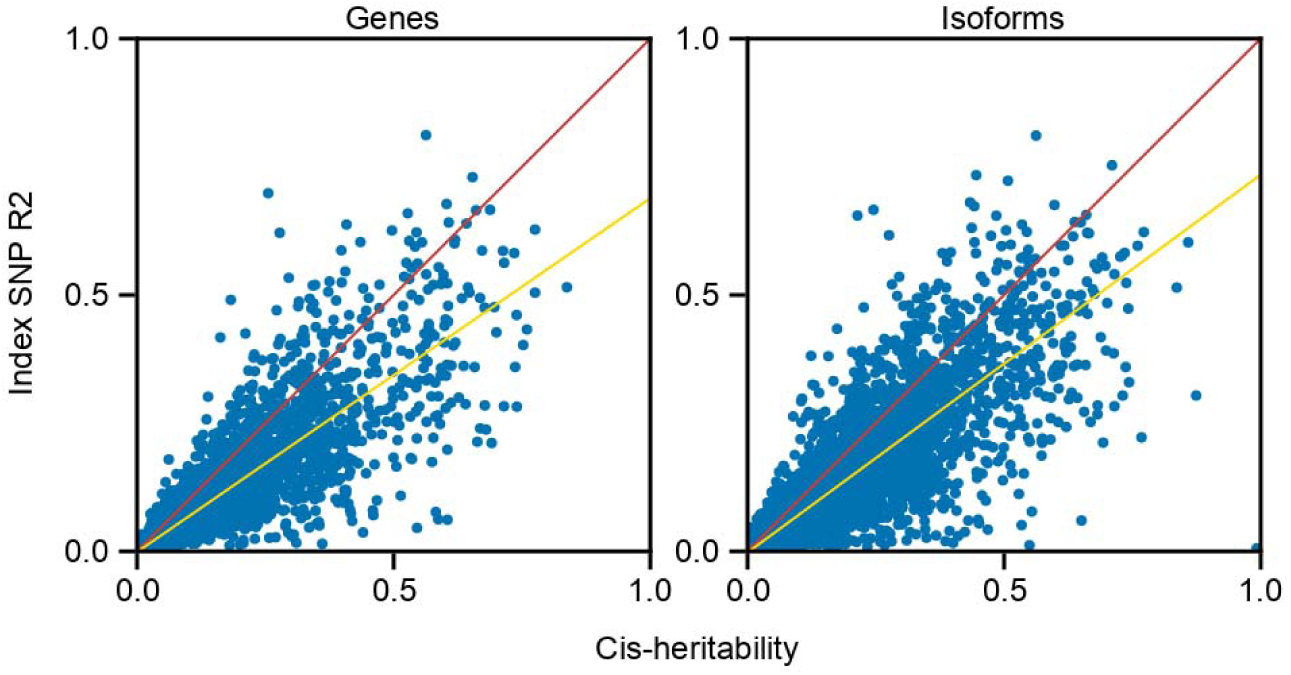
Sparsity of *cis*-SNP effects. Left, comparison between *h*^2^_cis_ estimates and variance explained by top associated eQTL SNPs for heritable genes. Right, the same comparison but for heritable isoforms.

**Figure S6:**
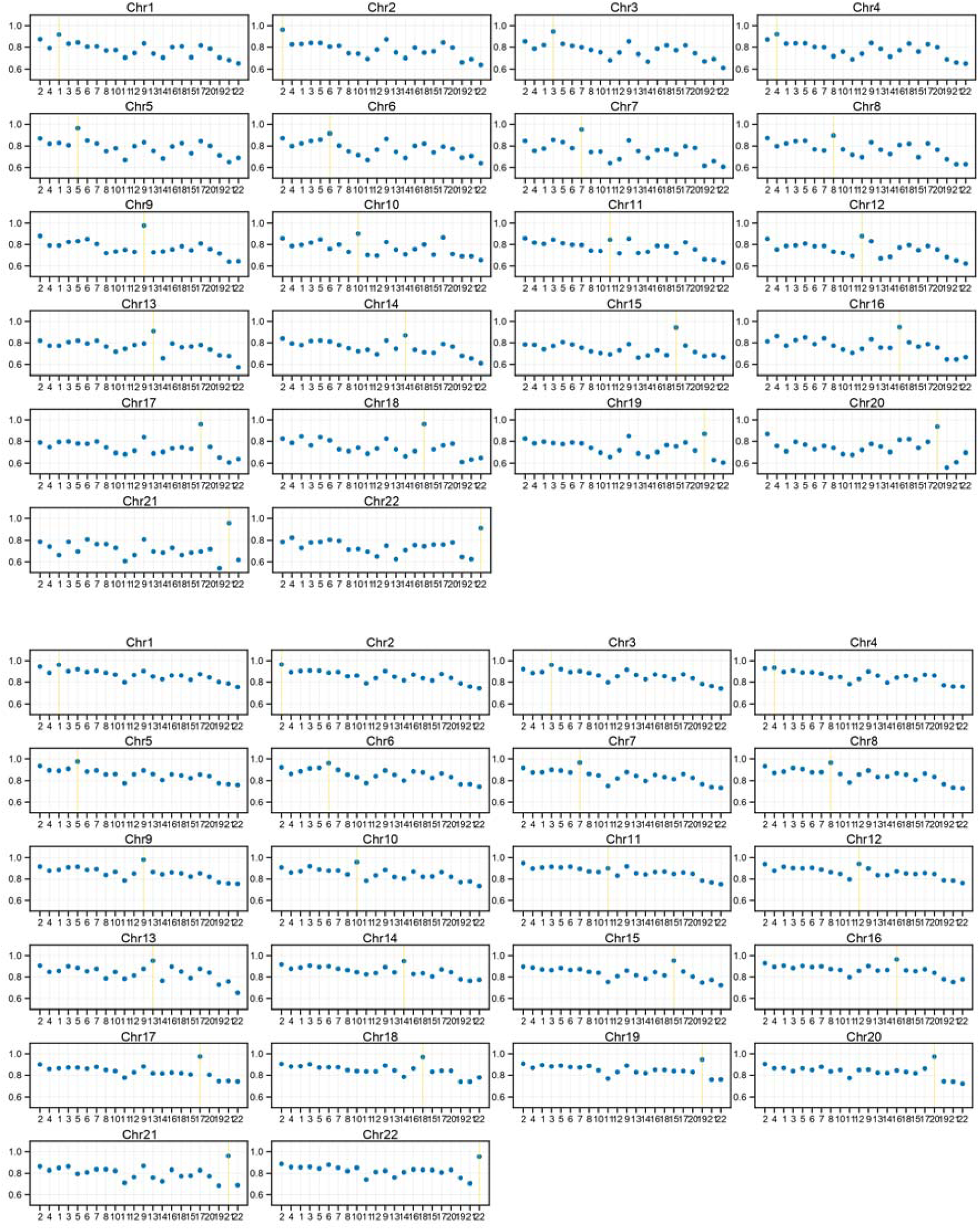
Polygenicity of *trans*-SNP effects. We fit a univariate penalized variance components model with 23 variance components parameters that capture genetic effects from each autosome and lasso penalty. Top, shown are results for genes. Bottom, shown are results for isoforms. For each chromosome, we focus on heritable genes and isoforms (*P* < 0.05) that reside in that chromosome, and for these features, we calculate the number of times each chromosome appeared in the solution path^22^. The chromosome the feature is from almost always appeared in the solution path (i.e. yellow lines have the highest proportions), while the rest of the chromosomes appeared in proportion to their number of SNPs. Chromosomes are listed in descending order of the number of SNPs on the x-axis.

**Figure S7:**
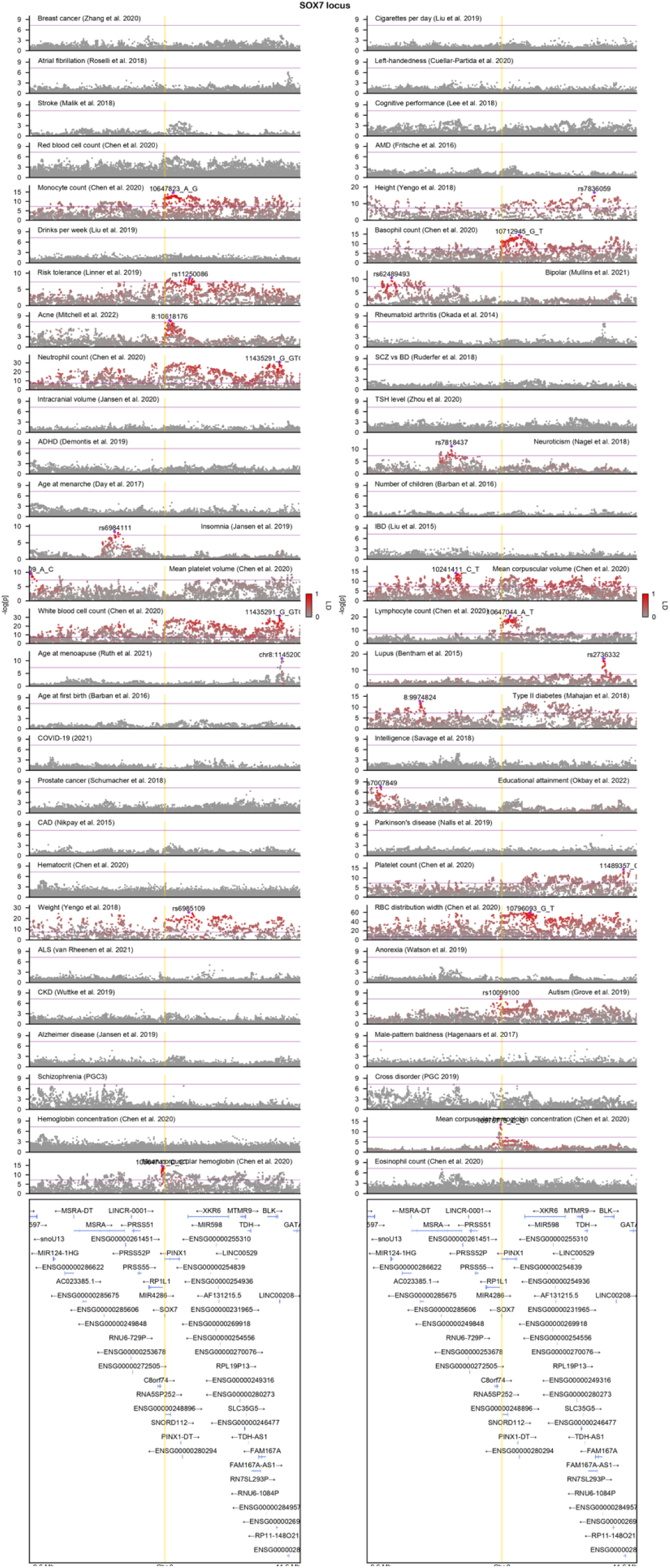
A close look at the ASD GWAS locus in chromosome 8. GWAS results for 56 complex phenotypes are shown, which span autoimmune, endocrine, psychiatric, cardiovascular disorders, and cancer. Index SNPs for phenotypes harboring GWAS hits are labeled and corresponding LD between other SNPs are displayed with the intensity of red color. Purple line denotes genome-wide significance (*P* = 5 × 10^−8^), and yellow lines denote gene start and end sites for *SOX7* gene. LD is calculated with individuals of European ancestry in the 1000 Genomes Project reference panel. ADHD (attention-deficit/hyperactivity disorder), ALS (amyotrophic lateral sclerosis), AMD (age-related macular degeneration), BD (bipolar disorder), CAD (coronary artery disease), CKD (chronic kidney disease), IBD (inflammatory bowel disease), RBC (red blood cell), SCZ (schizophrenia).

**Figure S8:**
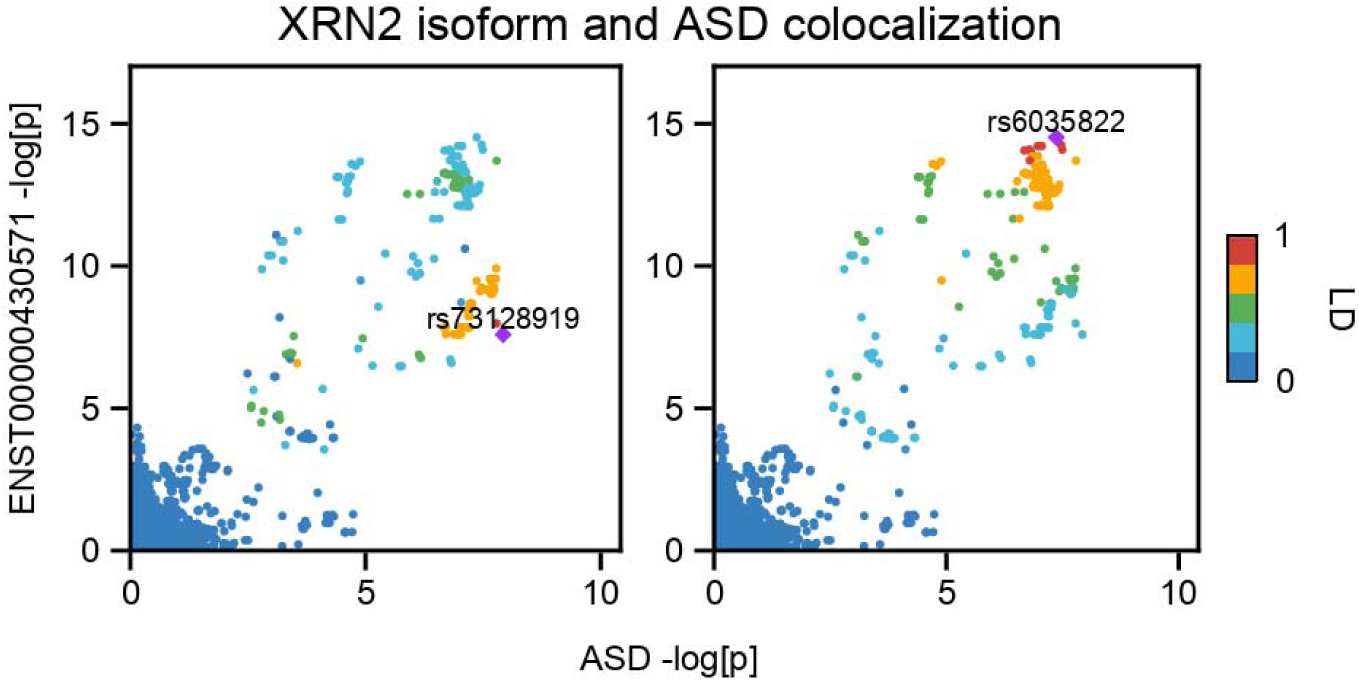
Colocalization between *XRN2* isoform-level eQTL and ASD GWAS results. Left, LD is colored with respect to the index SNP for ASD GWAS. Right, LD is colored with respect to the index SNP for ENST00000430571 eQTL. LD is calculated with individuals of European ancestry in PsychENCODE.

**Figure S9:**
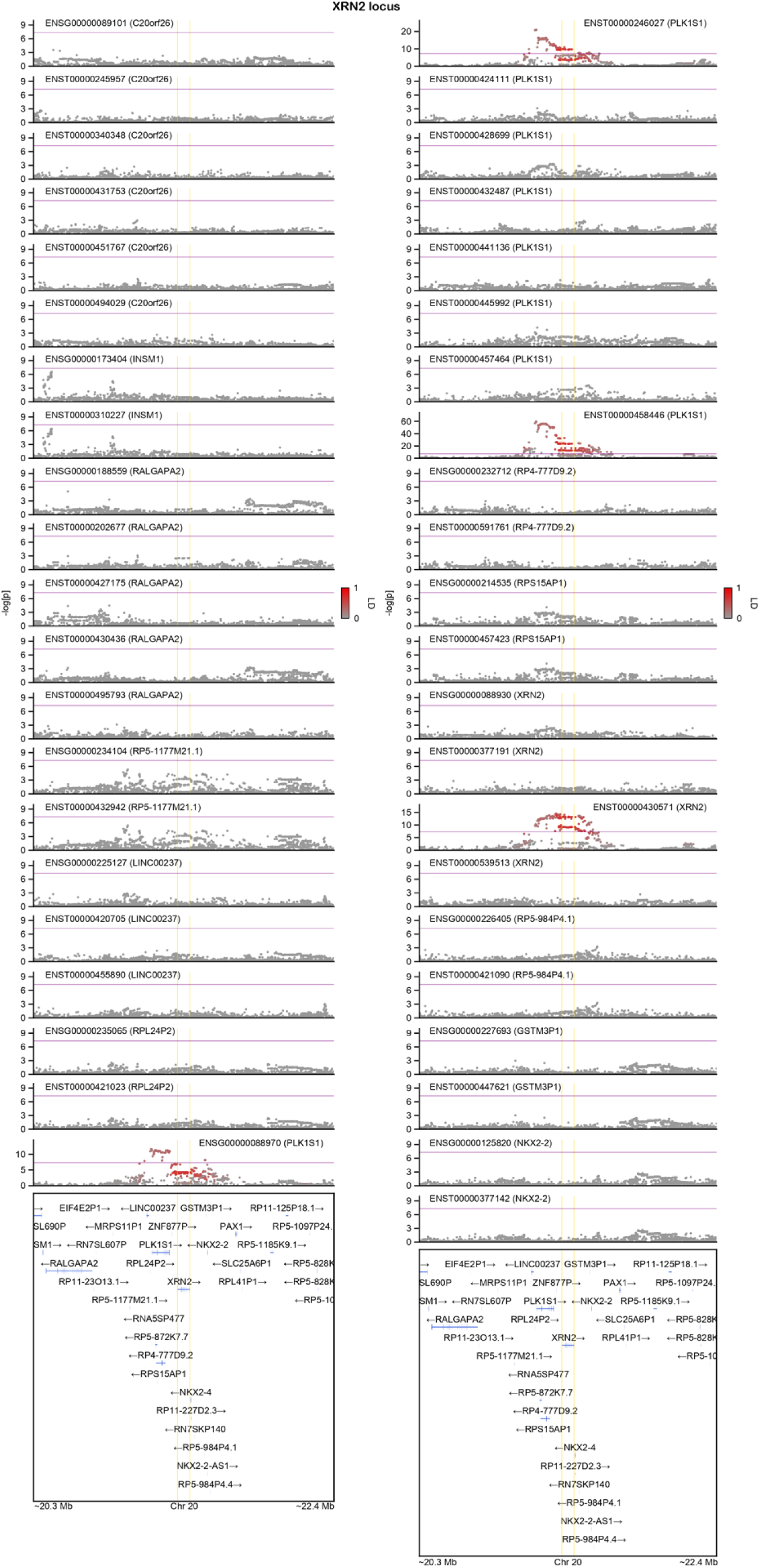
Absence of colocalization for all other features within the *XRN2* locus for the ASD GWAS signal. Shown is LocusZoom of eQTL signals for both gene- and isoform-level expression for all features within ±1 Mb window of (collapsed) gene start and end sites for *XRN2* gene. Gene names are shown in parentheses. LD is colored with respect to the index SNP for ASD GWAS (rs910805). LD is calculated with individuals of European ancestry in the 1000 Genomes Project reference panel.

**Figure S10:**
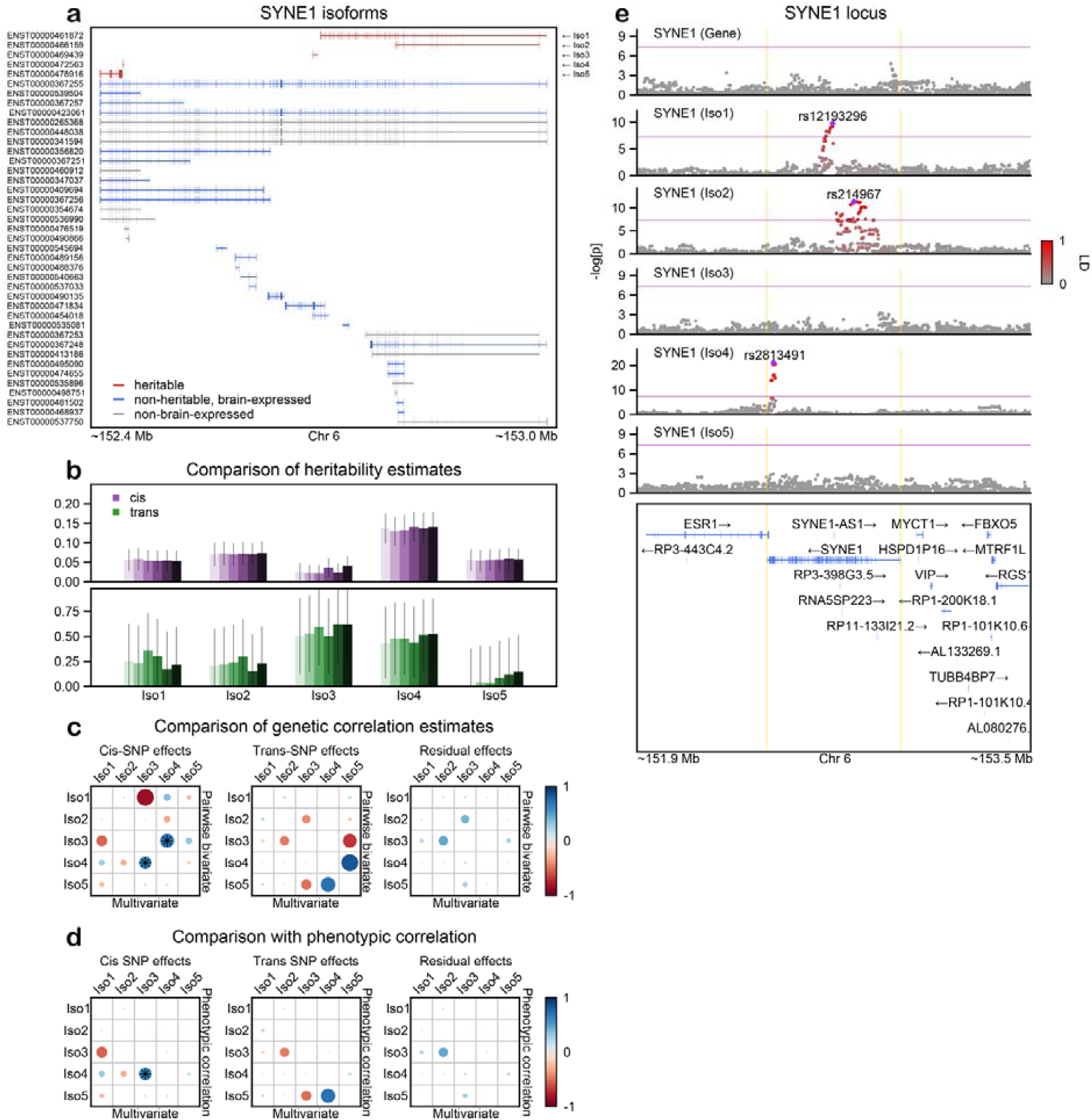
*h*^2^_SNP_, *r*_g_, and *cis*-eQTL results for *SYNE1* gene. Follows the same outline as **Figure 3** in the main text.

**Figure S11:**
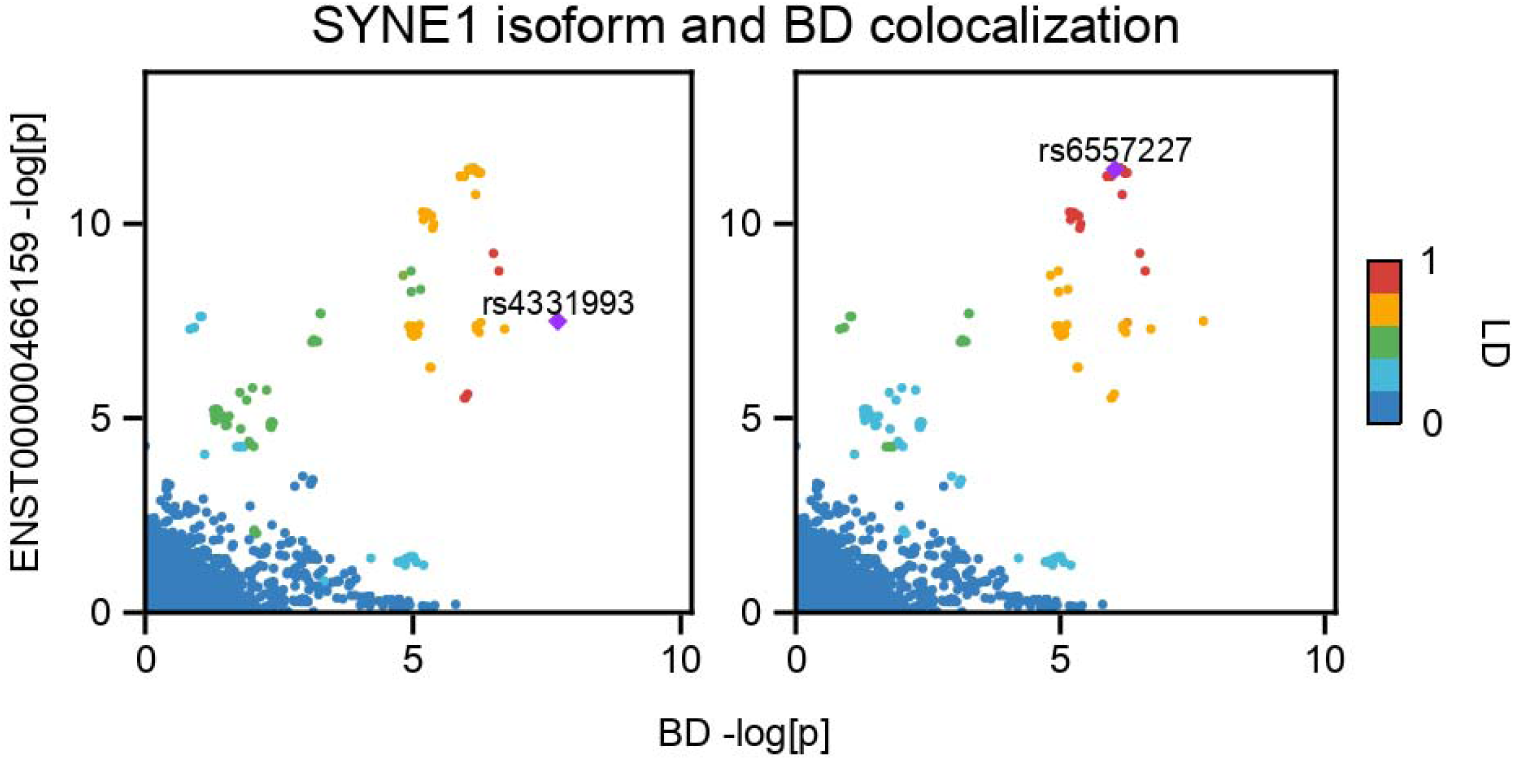
Colocalization between *SYNE1* isoform-level eQTL and BD GWAS results. Left, LD is colored with respect to the index SNP for BD GWAS. Right, LD is colored with respect to the index SNP for ENST00000466159 eQTL. LD is calculated with individuals of European ancestry in PsychENCODE.

**Figure S12:**
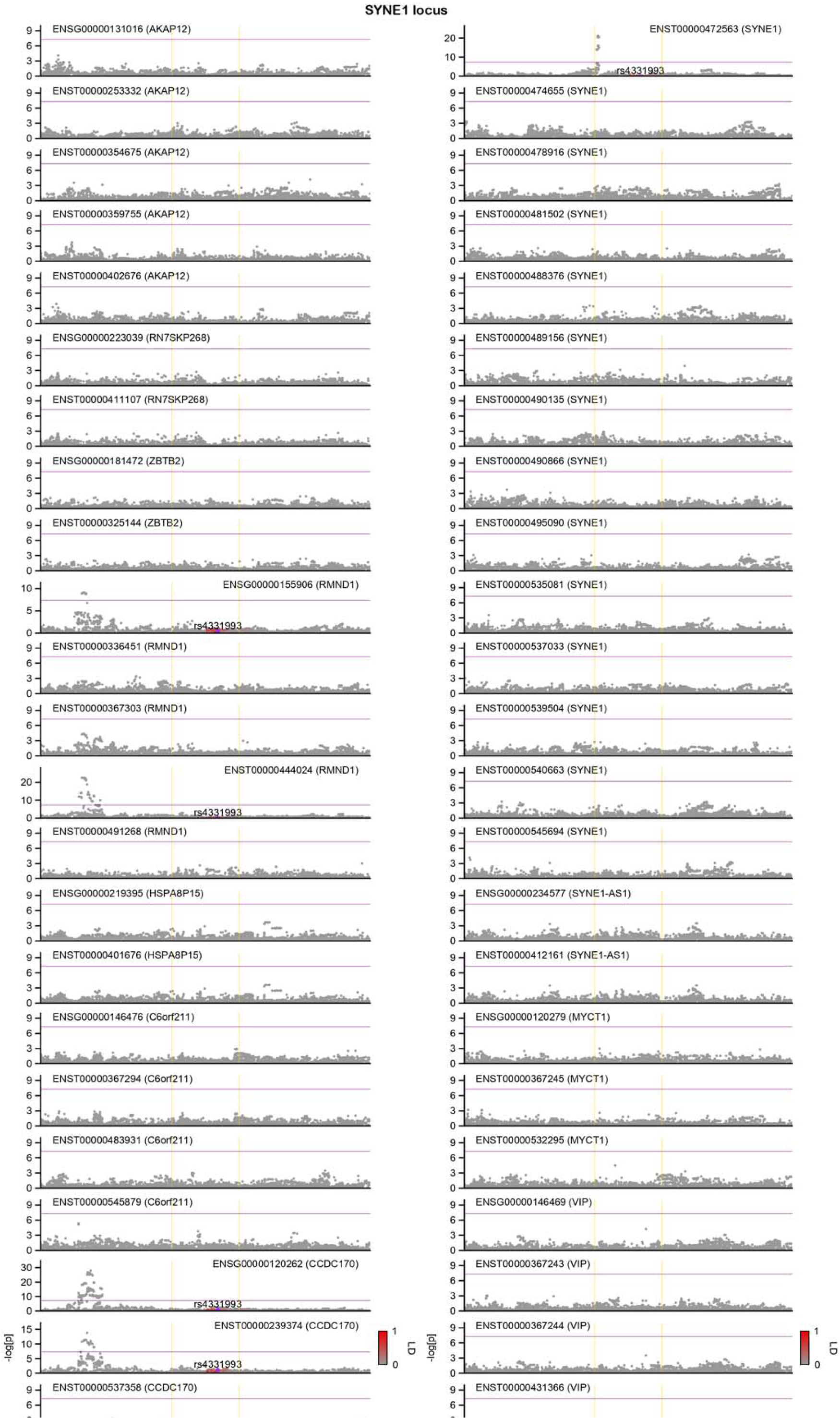
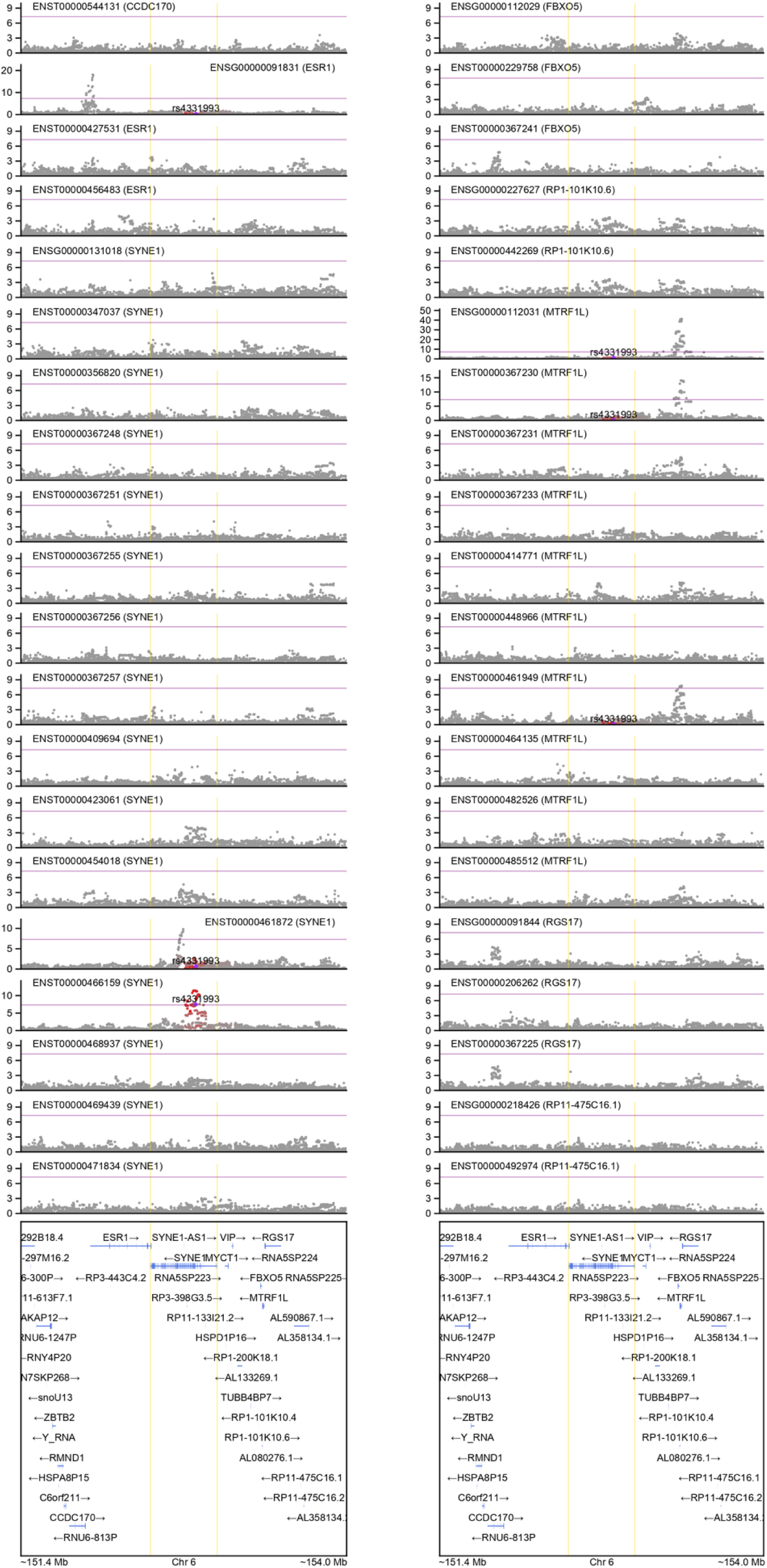
Absence of colocalization for all other features within the *SYNE1* locus for the BD GWAS signal. Shown is LocusZoom of eQTL signals for both gene- and isoform-level expression for all features within ±1 Mb window of (collapsed) gene start and end sites for *SYNE1* gene. Gene names are shown in parentheses. LD is colored with respect to the index SNP for BD GWAS (rs4331993). LD is calculated with individuals of European ancestry in the 1000 Genomes Project reference panel.

**Figure S13:**
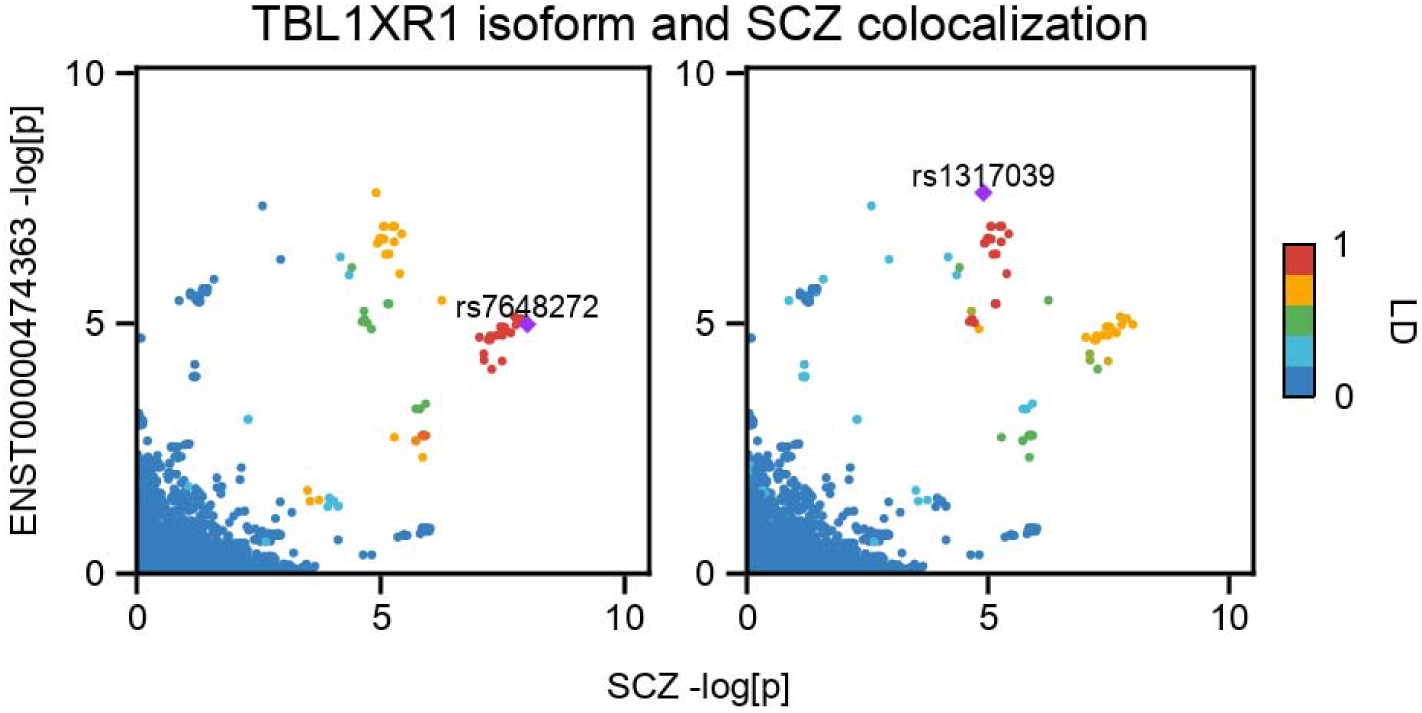
Colocalization between *TBL1XR1* isoform-level eQTL and SCZ GWAS results. Left, LD is colored with respect to the index SNP for SCZ GWAS. Right, LD is colored with respect to the index SNP for ENST00000474363 eQTL. LD is calculated with individuals of European ancestry in PsychENCODE.

**Figure S14:**
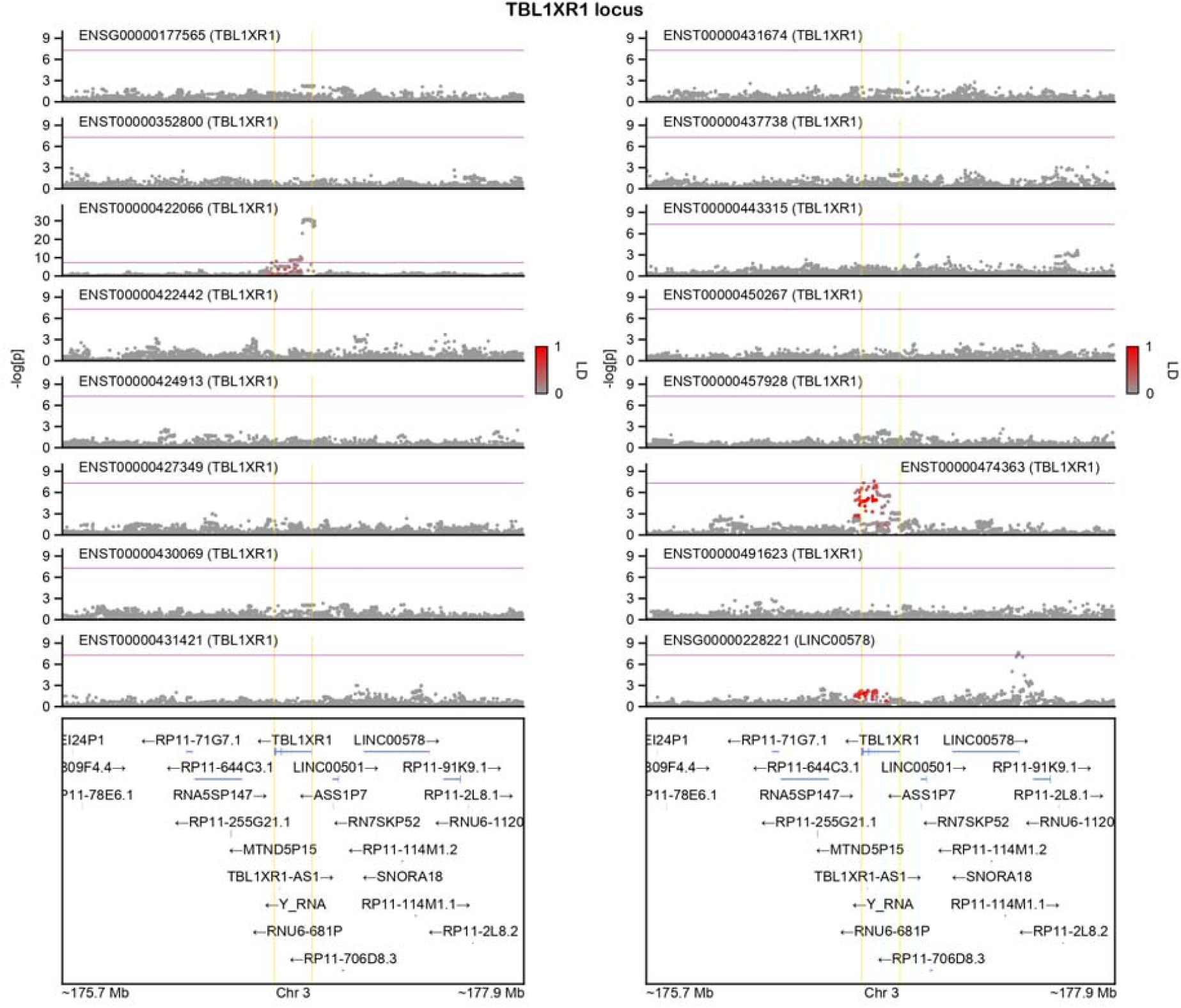
Absence of colocalization for all other features within the *TBL1XR1* locus for the SCZ GWAS signal. Shown is LocusZoom of eQTL signals for both gene- and isoform-level expression for all features within ±1 Mb window of (collapsed) gene start and end sites for *TBL1XR1* gene. Gene names are shown in parentheses. LD is colored with respect to the index SNP for SCZ GWAS (rs7609876). LD is calculated with individuals of European ancestry in the 1000 Genomes Project reference panel.

**Figure S15:**
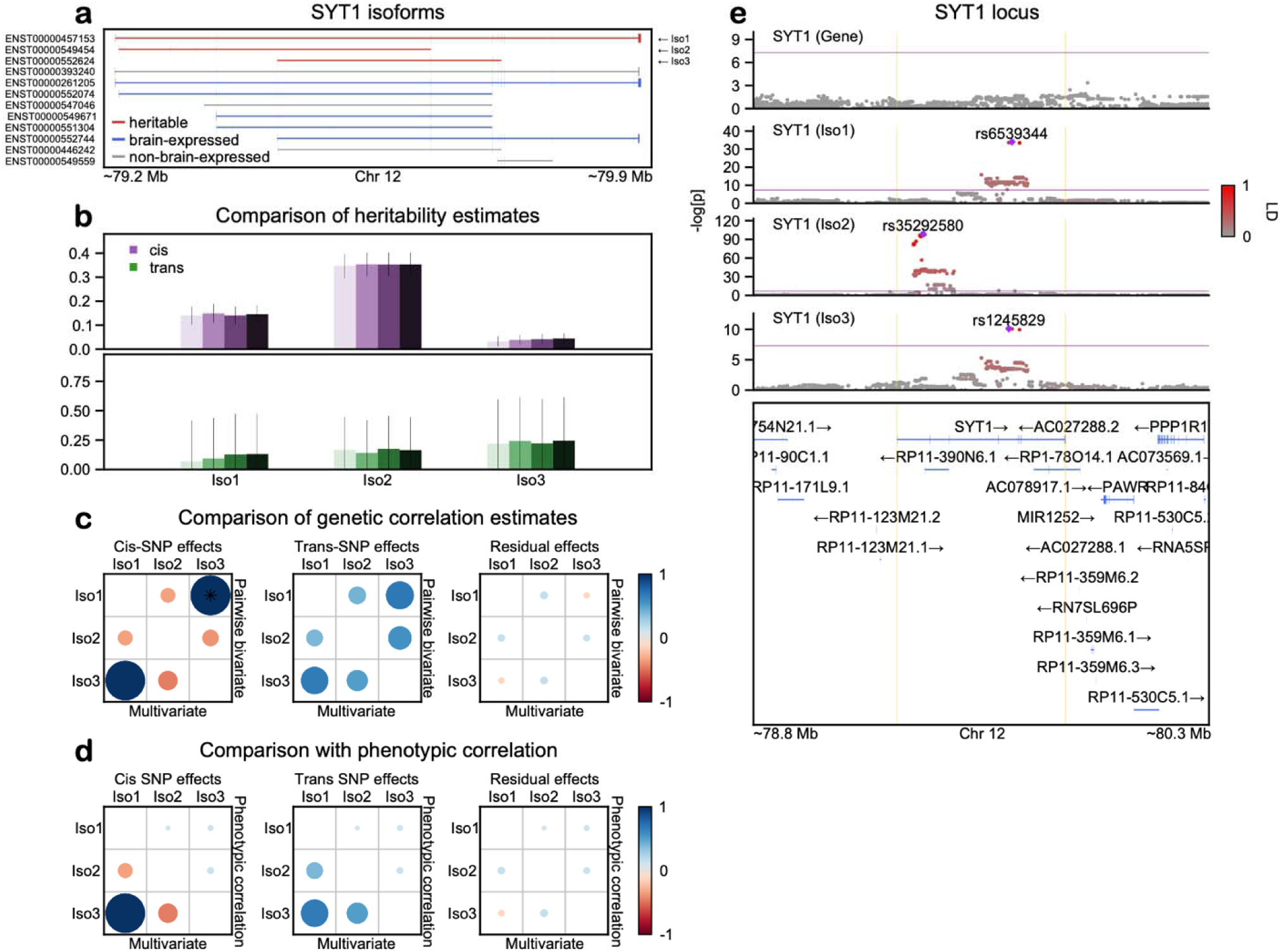
*h*^2^_SNP_, *r*_g_, and *cis*-eQTL results for *SYT1* gene. Follows the same outline as **Figure 3** in the main text.

**Figure S16:**
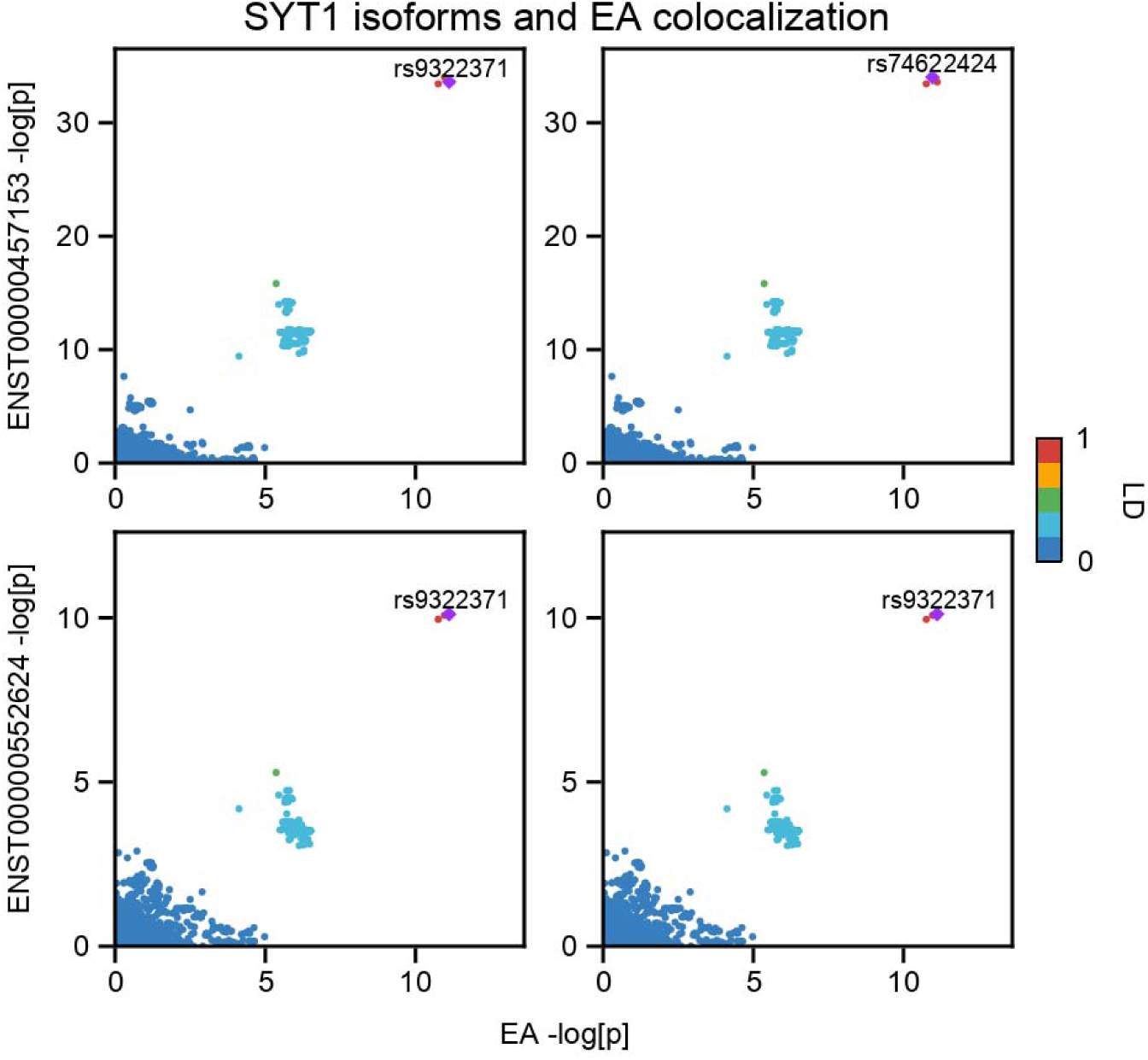
Colocalization between *SYT1* isoform-level eQTL and EA GWAS results. Left, LD is colored with respect to the index SNP for EA GWAS. Right, LD is colored with respect to the index SNP for corresponding isoform-level eQTL. LD is calculated with individuals of European ancestry in PsychENCODE.

**Figure S17:**
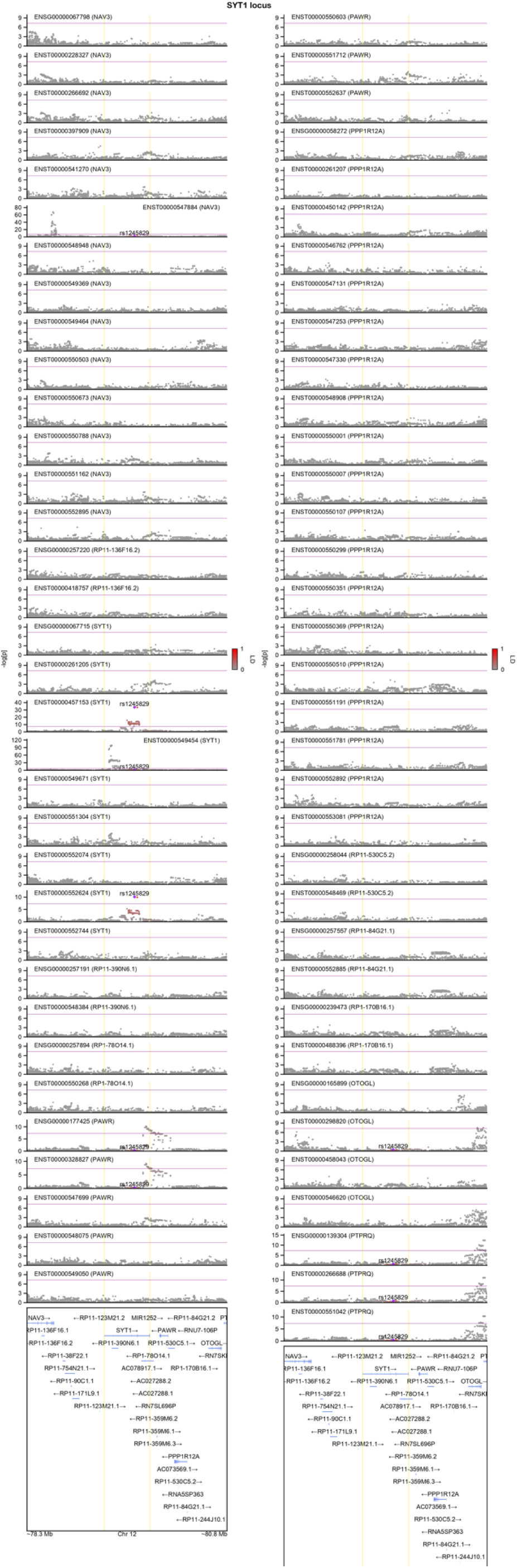
Absence of colocalization for all other features within the *SYT1* locus for the EA GWAS signal. Shown is LocusZoom of eQTL signals for both gene- and isoform-level expression for all features within ±1 Mb window of (collapsed) gene start and end sites for *SYT1* gene. Gene names are shown in parentheses. LD is colored with respect to the index SNP for EA GWAS (rs1245829). LD is calculated with individuals of European ancestry in the 1000 Genomes Project reference panel.

**Figure S18:**
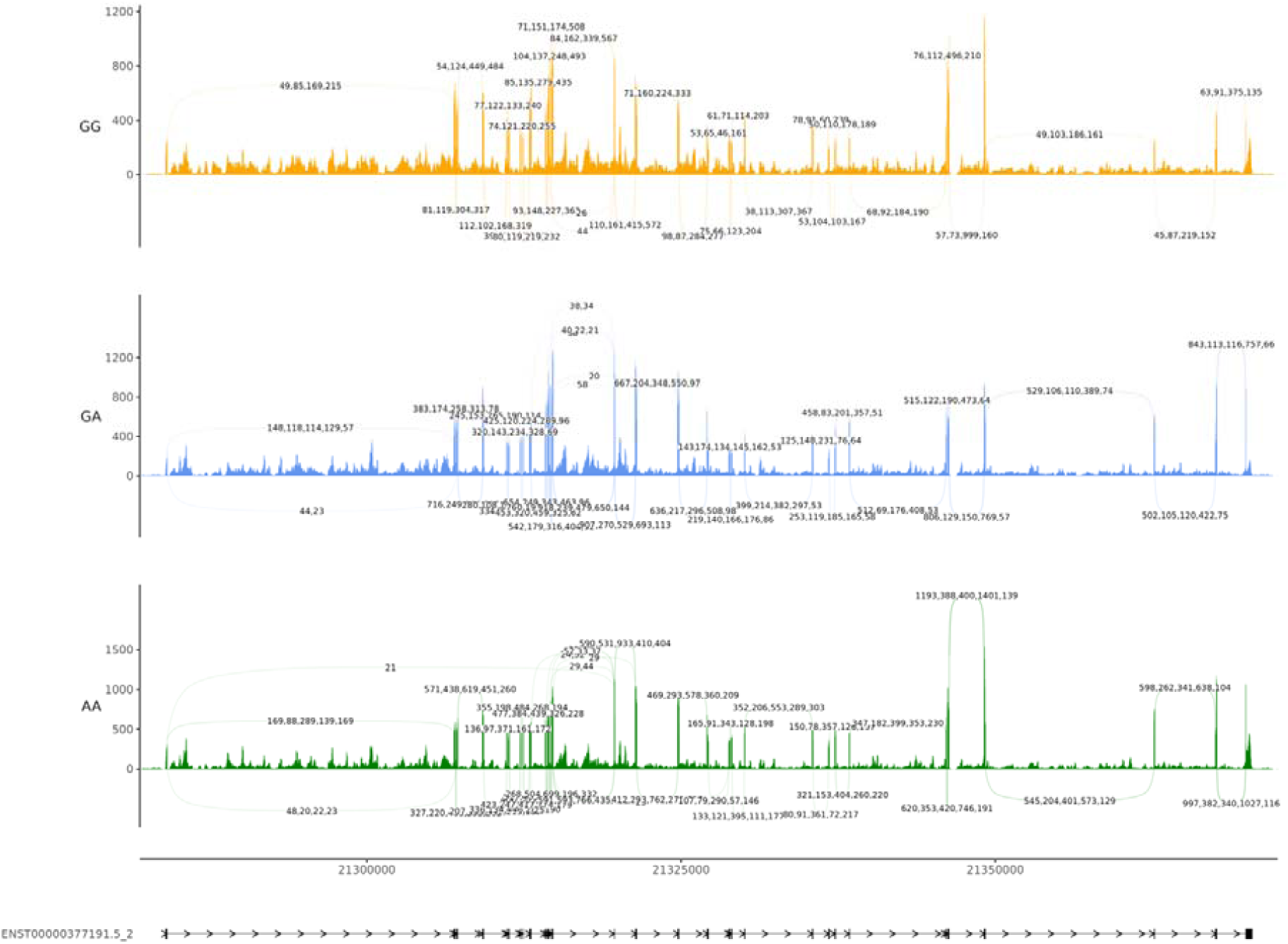
Local splicing events in the *XRN2* gene body across rs910805 genotypes. Five RNA-seq samples^47^ with approximately equal library (or read) depths are plotted for each rs910805 genotype. As above, the increase in the number of 1^st^ to 3^rd^ exon junction reads in the 5’ end with respect to the A major allele was subtle. This plot was generated with ggsashimi GitHub repository (https://github.com/guigolab/ggsashimi).

**Figure S19:**
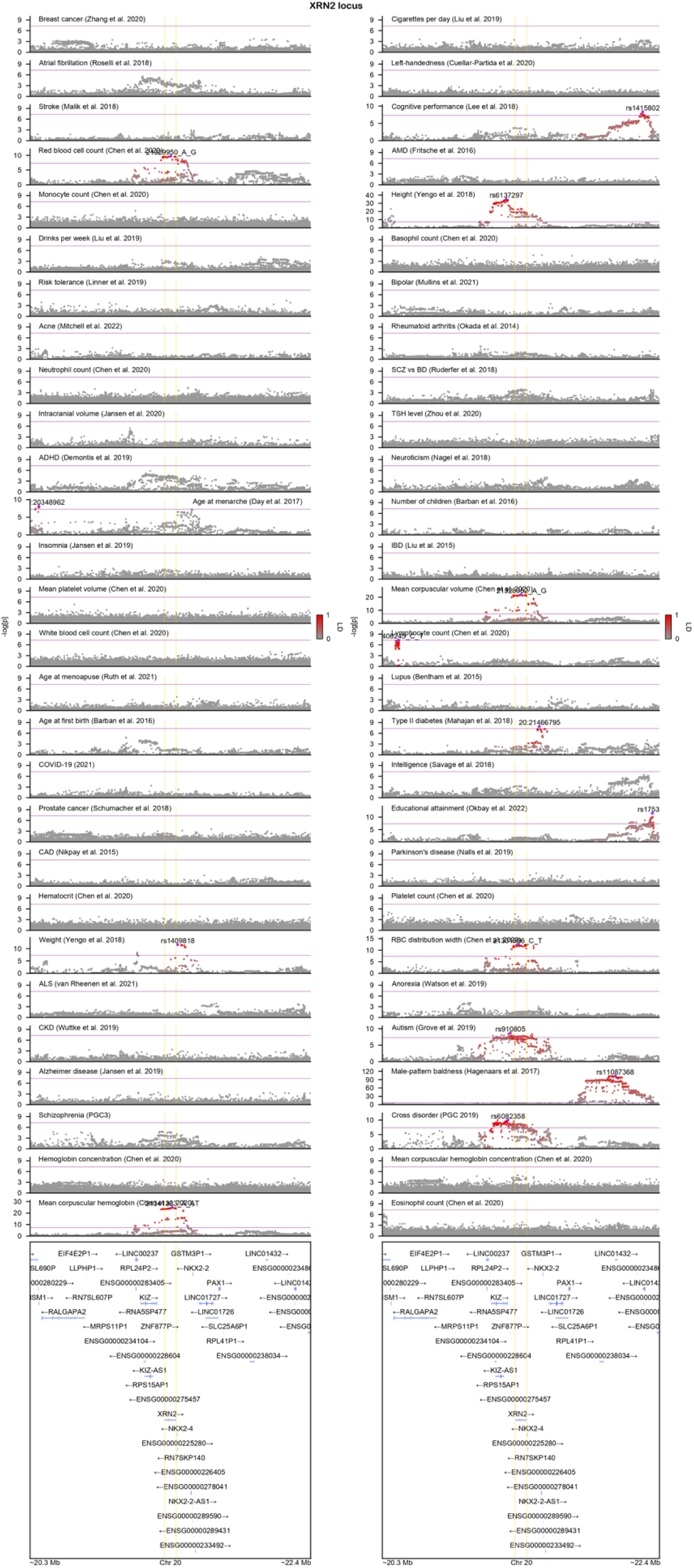
A close look at the pleiotropic *XRN2* locus. GWAS results for 56 complex phenotypes are shown, which span autoimmune, endocrine, psychiatric, cardiovascular disorders, and cancer. Index SNPs for phenotypes harboring GWAS hits are labeled and corresponding LD between other SNPs are displayed with the intensity of red color. Purple line denotes genome-wide significance (*P* = 5 × 10^−8^), and yellow lines denote gene start and end sites for *XRN2* gene. LD is calculated with individuals of European ancestry in the 1000 Genomes Project reference panel. ADHD (attention-deficit/hyperactivity disorder), ALS (amyotrophic lateral sclerosis), AMD (age-related macular degeneration), BD (bipolar disorder), CAD (coronary artery disease), CKD (chronic kidney disease), IBD (inflammatory bowel disease), RBC (red blood cell), SCZ (schizophrenia).

